# Blood donors as sentinels for genomic surveillance of West Nile virus in Germany (2020–2024) using a sensitive amplicon-based sequencing approach

**DOI:** 10.1101/2025.06.24.25329984

**Authors:** Gábor Endre Tóth, Marike Petersen, Francois Chevenet, Marcy Sikora, Alexandru Tomazatos, Alexandra Bialonski, Heike Baum, Balázs Horváth, Padet Siriyasatien, Anna Heitmann, Stephanie Jansen, Ruth Offergeld, Raskit Lachmann, Michael Schmidt, Jonas Schmidt-Chanasit, Dániel Cadar

## Abstract

West Nile virus (WNV) has emerged as a public health concern in Germany since its first detection in 2018, with evidence of expanding geographic spread. Genomic surveillance is critical for tracking viral evolution, identifying introductions, and monitoring local transmission. However, genome recovery from low-viremia samples such as those obtained through blood donor screening remains technically challenging. To develop and validate a sensitive amplicon-based sequencing protocol optimized for WNV lineage 2 and apply it to low-titer samples to support genomic surveillance in Germany. A novel primer scheme was designed for WNV lineage 2 and applied to 43 nucleic acid testing (NAT)-positive blood donor samples collected between 2020 and 2024. Amplicon-based sequencing performance was benchmarked against metagenomic next-generation sequencing (mNGS). Recovered genomes were subjected to phylogenomic analysis to assess viral diversity and transmission dynamics. The amplicon protocol enabled genome recovery (>70% coverage) from samples with viral loads as low as ∼10¹ RNA copies/µL, outperforming metagenomic NGS (mNGS). Of the 43 samples, 30 yielded complete or near-complete genomes. Six distinct WNV subclades (2A–2F), including German strains, were identified, indicating multiple introductions into Germany from Central Europe. Subclade 2F emerged as the dominant and widely distributed group. Berlin, Brandenburg, Saxony, and Saxony-Anhalt were identified as persistent transmission hubs. This study highlights blood donors as valuable sentinels for WNV genomic surveillance. The validated amplicon-based sequencing approach enables sensitive, scalable genome recovery from low-viremia samples, and when integrated with routine blood donor screening, provides a robust framework for early detection, transmission dynamics, and public health preparedness.

## Introduction

West Nile virus (WNV) or *Orthoflavivirus nilense* is a mosquito-borne orthoflavivirus that has become an emerging public health threat in the recent decade and a significant cause of viral encephalitis in humans and horses in many temperate regions [1]. Maintained in an enzootic transmission cycle between birds and *Culex* mosquitoes, WNV occasionally spills over into humans and other mammals, which serve as dead-end hosts [2]. Most human infections are asymptomatic, while ∼20% develop West Nile fever (WNF) and less than 1% progress to West Nile neuroinvasive disease (WNND), particularly elderly or immunocompromised individuals [1,3]. Over the past decade, Europe has witnessed a notable increase in WNV activity and an expanding geographic range of transmission. Following its establishment in Southeastern Europe in the late 1990s, the virus has expanded northward, facilitated by climate change, land use patterns, and migratory bird movements [4,5]. A record number of human cases in 2018 (n=2,083) and another resurgence in 2022, with over 1,300 cases reported across the EU/EEA [6,7]. In Germany, the first local WNVbeast detections occurred in 2018 in animals, with confirmed endemic transmission in birds, horses, and humans in subsequent years, particularly in Berlin and the Eastern federal states [8]. Surveillance has shown that Germany’s WNV activity stems from WNV lineage 2 strains introduced from Central Europe [5]. Timely diagnosis and surveillance of WNV are challenged by the typically low and transient viremia in human infections. Nucleic acid amplification tests (NATs) are sensitive during the brief viraemic phase but may fail to detect infections later in the course of disease [9]. Serological testing is useful but limited by cross-reactivity with other orthoflaviviruses and by the prolonged persistence of IgM antibodies, as observed in patients with neuroinvasive WNV infection, both of which complicate result interpretation [10]. These diagnostic challenges also impact the ability to recover viral genomes from clinical samples for molecular epidemiology. Genomic surveillance has become an essential component of arbovirus monitoring, as genome (>70%) sequencing enables detailed tracking of viral lineages, mutations, and transmission pathways [5,11]. While metagenomic next-generation sequencing (mNGS) directly from specimens can detect WNV without prior knowledge, in practice, it often lacks sufficient sensitivity when viral RNA levels are low, or background host nucleic acid is abundant. Capture-based enrichment techniques offer higher sensitivity but are resource-intensive. In contrast, amplicon-based sequencing using virus-specific primers offers a practical and highly sensitive alternative. Several amplicon-based protocols have recently been developed for WNV genome recovery directly from clinical, veterinary, and mosquito samples [12–18]. However, existing methods demonstrate sensitivity limitations: complete or near-complete genome recovery becomes difficult in samples with high cycle threshold (Ct) values (>31,5), a common status in asymptomatic infections and blood donors. These limitations are particularly problematic for blood donor surveillance, where early detection is critical but viral loads are typically very low. Blood donor screening represents a unique and valuable system for surveillance of WNV. Donations are systematically collected, geographically widespread, and subjected to highly sensitive NAT testing, making blood donors an effective early-warning group for WNV circulation. However, the low viral RNA concentrations in these samples have limitations in successful genome recovery using existing methods, restricting phylogenetic and epidemiology insights into circulating strains [19,20]. To address this gap, we developed a novel, highly sensitive amplicon-based WNV lineage 2 whole-genome sequencing protocol tailored for surveillance in samples with low viral load. In this study, we validate the newly developed method using blood donor samples collected during the 2020–2024 WNV transmission seasons in Germany and conduct a comprehensive genomic, epidemiological, and evolutionary analysis of the current WNV activity in Germany.

## Methods

### WNV-positive blood donor samples

In Germany, screening of blood donations for WNV using NAT is required during the transmission season since 2000, as stipulated in a regulatory notification by the Paul-Ehrlich-Institute under §28 of the Medicinal Products Act (Arzneimittelgesetz, AMG). This directive aims to minimize the risk of WNV transmission through blood components and stem cell preparations. Alternatively, few blood establishments continue to apply a precautionary deferral of donors who have spent at least two days in endemic areas [19]. Roughly 2.0-2.4 million donations were screened yearly between 2020 and 2024. A total of 43 serum samples from blood donors that tested positive for WNV by NAT screening and were subsequently confirmed as WNV-positive via qRT-PCR/mNGS in our laboratory were included in this study. These samples were collected between 2020 and 2024 from various blood establishments across Germany and accounted for the majority of confirmed WNV-positive donations. Sample metadata are provided in Table S1.

### Development of an amplicon-based protocol for WNV genome sequencing

We collected all complete (>80%) WNV lineage 2 sequences from NCBI GenBank (2023-12-01). A total of 495 genomes were selected, and primer schemes were generated using PrimalScheme v1.4.1 [17]. The scheme targets approximately 10,815 bp of the WNV genome with 59 overlapping amplicons. To capture known lineage 2 diversity, alternative primers were included, resulting in two primer pools with 189 primers total (Pool_1: 99, Pool_2: 90). Amplicon sizes ranged from 252 to 280 bp (mean: 268.3 bp, SD ±5.8) (Figure S1). Primer details, source sequences, as well as a detailed description of the whole protocol, are available at protocols.io (https://www.protocols.io/view/west-nile-virus-orthoflavivirus-nilense-lineage-2-q26g71q98gwz/v1). Before implementation, three *in vitro* WNV isolates (UG37, B956, BNI-129) were sequenced in serial dilution to optimize reaction conditions and optimize the protocol.

### Metagenomic next-generation sequencing of WNV positive blood donors

WNV-positive blood donor samples were also subjected to unbiased metagenomic next-generation sequencing (mNGS) to further investigate NAT-reactive cases. RNA extracted and purified from plasma or serum samples was processed using an in-house mNGS pipeline established for virus discovery [21] and sequenced on a NextSeq 2000 platform (Illumina).

### Genome assembly and data acquisition

Raw reads generated by the amplicon-based approach were checked using FastQC v0.12.1 [22] for quality assessment. Paired-end reads were merged with BBMerge v38.84 [23], and size-selected between 240–290 bp to match expected amplicon lengths. Primer trimming and quality filtering were performed using BBDuk v38.84. Clean reads were aligned to the WNV reference genome (GenBank: MN794937.1) using Bowtie2 v2.4.5, [24] and consensus sequences were generated with SAMtools v1.18 [25]. Mapping accuracy was visually inspected using Geneious Prime v2024.0.7. The complete workflow is summarized in Figure S2. For comparative analyses, all publicly available nucleotide sequences of European WNV strains were retrieved from the NCBI Nucleotide database (https://www.ncbi.nlm.nih.gov) as of February 2025. The final compiled dataset consisted of 520 complete or near-complete genome sequences, spanning the years 2004 to 2024. Geospatial coordinates for all samples included in the dataset were standardized and integrated (Table S2).

### Phylodynamic reconstruction of the WNV spread pattern

To infer evolutionary timelines and characterize both cross-border and intra-national viral transmission patterns, we reconstructed time-scaled phylogenetic trees focusing on the dominant European WNV lineage 2a. Two analytical scenarios were considered: (i) a full European WNV dataset and (ii) a subset restricted to WNV transmission dynamics for Germany. Bayesian time-scaled phylogenies were reconstructed using BEAST v1.10.5 [26], applying an uncorrelated relaxed molecular clock model and a Skygrid coalescent prior. Model selection was informed by stepping-stone sampling. For the full dataset, MCMC chains were run for 400 million steps, and for the German subset, 200 million steps, with parameters sampled every 10,000 steps.

### Phylogeography and spatiotemporal dynamics of WNV in Germany

To reconstruct viral migration patterns in Germany, all publicly available complete or near-complete WNV sequences from both human and animal cases were included in the phylogeographic analyses, alongside the newly generated blood donor sequences from this study. We employed an asymmetric discrete trait phylogeographic model combined with the Bayesian Stochastic Search Variable Selection (BSSVS) procedure to identify statistically supported transition routes. In addition, a continuous phylogeographic model based on the Relaxed Random Walk (RRW) framework was implemented to visualize the spatial diffusion of WNV in continuous geographic space [27]. These analyses also provided estimates for the time to the most recent common ancestor (tMRCA). Maximum clade credibility (MCC) trees were generated and visualized using FigTree v1.4.1 (http://tree.bio.ed.ac.uk/software/figtree/). The spatiotemporal dynamics of WNV were further explored and visualized through EvoLaps 2 (www.evolaps.org), enabling advanced phylogeographic reconstruction and representation [28].

## Results

### Validation of the WNV amplicon-based sequencing protocol

For our initial validation, we first tested 3 in vitro WNV isolates (UG37, B956, BNI-129) and sequenced them across 10-fold serial dilutions. The corresponding Ct values ranged from 19.45 to 40.52, reflecting viral RNA concentrations from 667225 to 0.13 copies/µL. Sequencing yielded a mean depth of 309295 reads (SD ±66,297.3). A marked decline in genome recovery was noted at concentrations below 8.1 copies/µL. At higher input levels (>8.1 copies/µL), over 70% of the WNV genome from all three genetically diverse WNV isolates was recovered at ≥10× coverage (Figure S3; Table S3). To assess its performance, we validated the protocol using 43 WNV-positive blood donor serum samples. Ct values ranged from 26.92 to 43.63, corresponding to viral loads between 2,797.97 and 0.01 WNV RNA copies/µL (Figure 1a). The mean sequencing depth was 771,805.7 single reads per sample (SD ± 589,487.5) while the median was 561,238. Only 6 samples (13%) exceeded 1 million reads in total sequencing depth (Figure S4; Table S4). Altogether, 30 out of 43 blood donor samples were successfully recovered. All samples showed over 70% genome coverage up to a Ct value of 35.12, with two additional samples exceeding this threshold despite lower viral loads (Ct 35.62 and 36.35). We applied a generalized linear model (GLM) to estimate the WNV RNA copy number required to surpass the 70% genome recovery threshold. Based on our dataset, a concentration of 11.2 copies/μL was sufficient to recover usable genomes at ≥10× depth, while at ≥1× depth, the threshold decreased to 6.9 copies/μL (Figure 1b, Figure S4).

**Figure 1.**
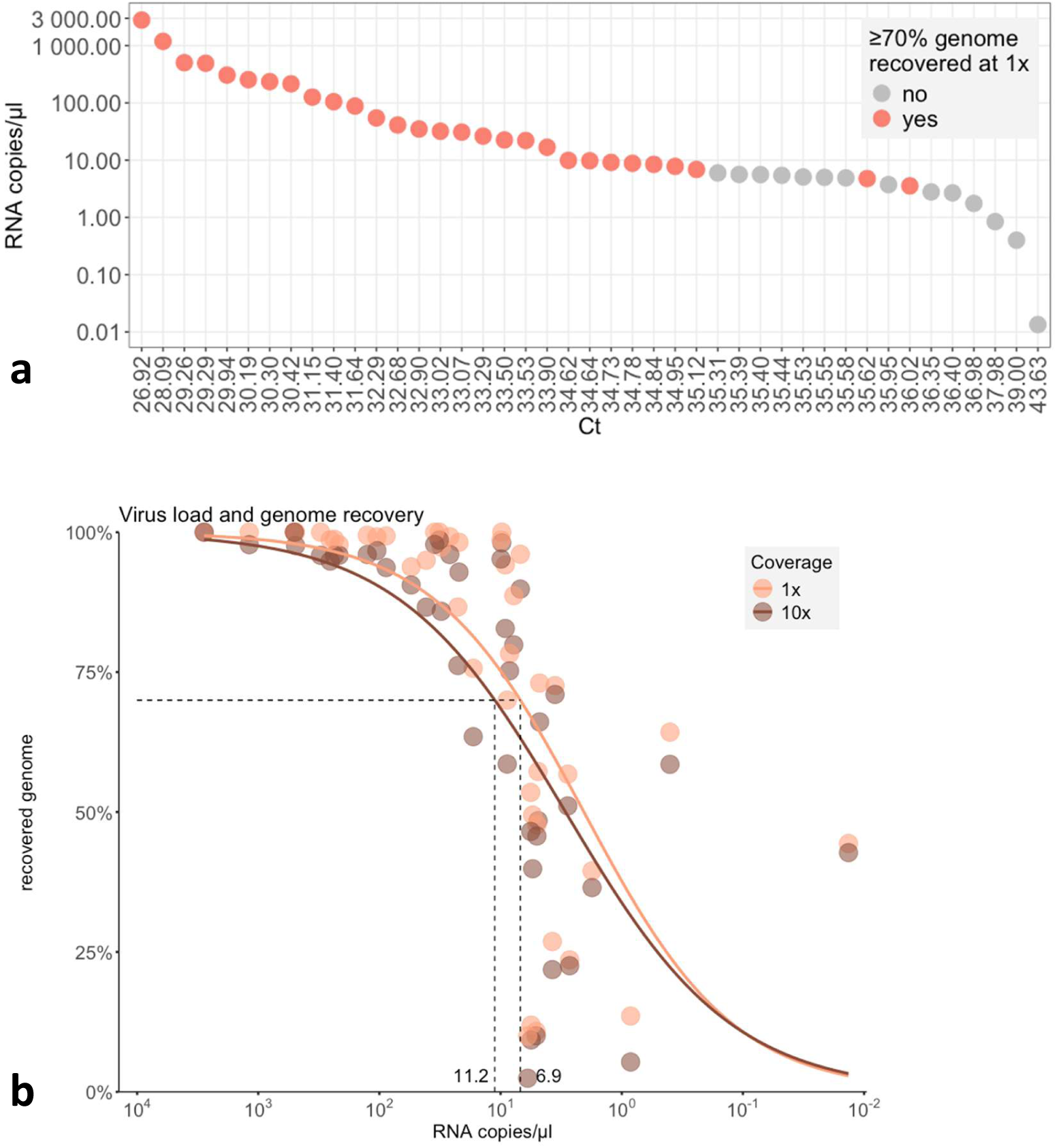
Performance of WNV amplicon-based sequencing across blood donor samples. **Panel a** assesses performance by ≥70% genome recovery at 1× coverage, while **Panel b** shows complete genome recovery at both 1× and 10× coverage. The x- and y-axes of **Panel a** display Ct values and viral RNA copies for each analyzed blood donor sample. Panel b presents viral RNA copies and percentage genome coverage at 1x and 10x read depth. The performance of the sequencing protocol was evaluated using R v4.5.0 and the packages dplyr v1.1.4, ggplot2 v3.5.2, readxl v1.4.5, scales v1.4.0, ggpubr v0.6.0, gggenes v0.5.1, gridExtra v2.3, and patchwork v1.3.0.

The efficiency of certain amplicons (e.g., amplicons 23 and 58) was reduced, likely due to chimeric read formation, which contributed to discrepancies in genome recovery between 1× and 10× coverage thresholds (Figure 1). While the proportion of WNV-specific reads generally correlated with viral concentration, a marked drop was observed at Ct values around 35. Moreover, several outliers deviated from this trend, underscoring the impact of sample quality on sequencing success (Figure S4, S5). Overall, these results indicate that our protocol performs reliably across a broad range of viral loads and sample conditions, achieving comparable efficiency to that observed during in vitro validation with serially diluted virus isolates.

### Metagenomics versus targeted sequencing for WNV genome recovery

We compared metagenomic and amplicon-based sequencing approaches for WNV genome recovery using the same panel of WNV-positive blood donor samples with varying viral loads. Both methods demonstrated high sensitivity, reliably detecting WNV across all samples. However, significant differences in genome recovery were observed. With the metagenomic approach, full genome recovery at 1× coverage was achieved from samples with ≥10² RNA copies/µL, and at 10× coverage from ≥10³ copies/µL (Figure S6). In contrast, the amplicon-based approach outperformed the metagenomics approach, enabling appropriate genome recovery at 1× coverage from less than 10 RNA copies/µL, and 10× from ∼10¹ copies/µL (Figure S6). These findings highlight the increased sensitivity and genome completeness of the targeted amplicon strategy, particularly for low-copy clinical specimens.

### Genomic characterization and phylogenetic placement of WNV from blood donors

A total of 43 West Nile virus-positive blood donor samples collected between 2020 and 2024 were characterized based on complete or partial genome sequences. These donors originated from both known endemic regions (e.g., Brandenburg, Saxony, Saxony-Anhalt) and newly affected federal states, including Schleswig-Holstein and Lower Saxony, reflecting the expanding geographic distribution of WNV circulation in Germany (Table S1; Figure 2). The genetic variation of German WNV strains across the viral genome was relatively heterogeneous, exhibiting at least 94 synonymous and 23 non-synonymous mutations (Figure S7). Despite this variation, the virus has maintained a highly conserved genome since its first detection in Germany in 2018. The genome-wide and gene-specific identity matrices were consistently greater than 99% at both the nucleotide and amino acid levels. Bayesian maximum clade credibility (MCC) phylogenetic analysis of the complete European WNV dataset revealed that all German strains, including those derived from blood donors, clustered within a single major clade of the WNV lineage 2 phylogeny (Figure S8). A more detailed analysis of this clade indicated that the German sequences could be classified into six distinct subclades, provisionally designated as subclades 2A–2F. Subclades 2A, 2D, 2E, and 2F exhibited monophyletic structures, suggesting single introduction events into Germany for these groups. In contrast, subclades 2B and 2C were polyphyletic, indicating that German strains within these groups evolved from multiple ancestral Austrian strains (Figure 3). Blood donor-derived WNV genomes were distributed across five of the six subclades, except for subclade 2B, from which no blood donor sequences were identified (Figure 3). Among the identified subclades, subclade 2F emerged as the dominant group in Germany, including the majority of recent WNV genomes sampled during the 2020-2024 mosquito seasons. This observation suggests that subclade 2F has undergone successful local expansion and may represent an established endemic variant within the German WNV population. Analysis of the complete polyprotein sequences identified subclade-defining and/or geographically associated point mutations (Figure S7). Notably, all members of subclade 2F, which has emerged as the dominant group in Germany, harbour a unique amino acid substitution (K609R) within the helicase domain of the NS3 protein (Figure S7).

**Figure 2.**
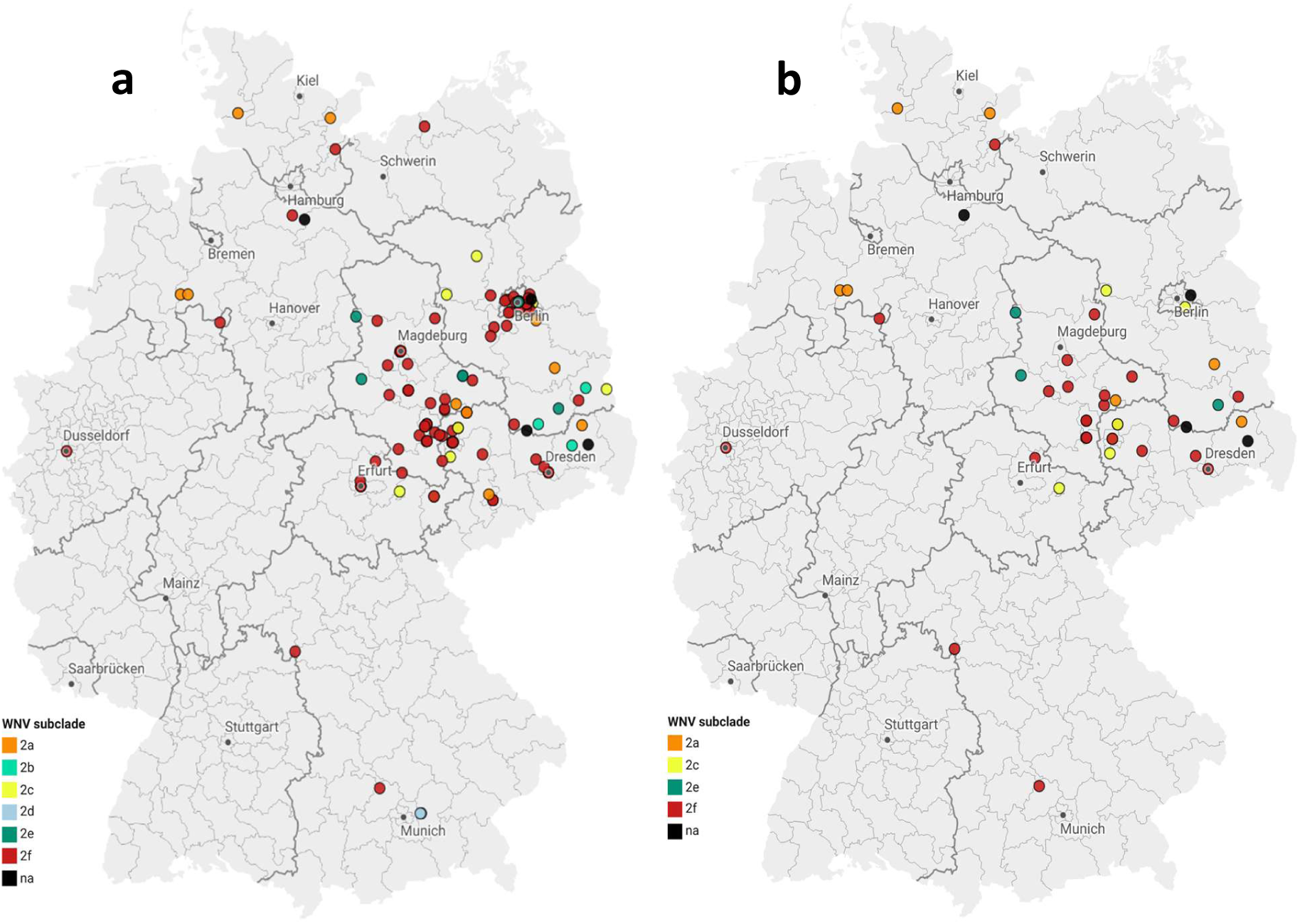
Geographical distribution of West Nile virus in Germany (2019–2024) **Panel a** shows the location of all WNV genome sequences from Germany that are publicly available in GenBank, covering both human and animal cases. This comprises previously published sequences (from birds, horses, and symptomatic human infections) as well as the blood donor sequences generated in our current study, while **panel b** displays locations of WNV-positive blood donors identified and sequenced in the present study. Coloured circles denote the genetic clustering of detected WNV strains within subclades 2A–2F, based on phylogenetic reconstruction. Map created with Datawrapper.

**Figure 3.**
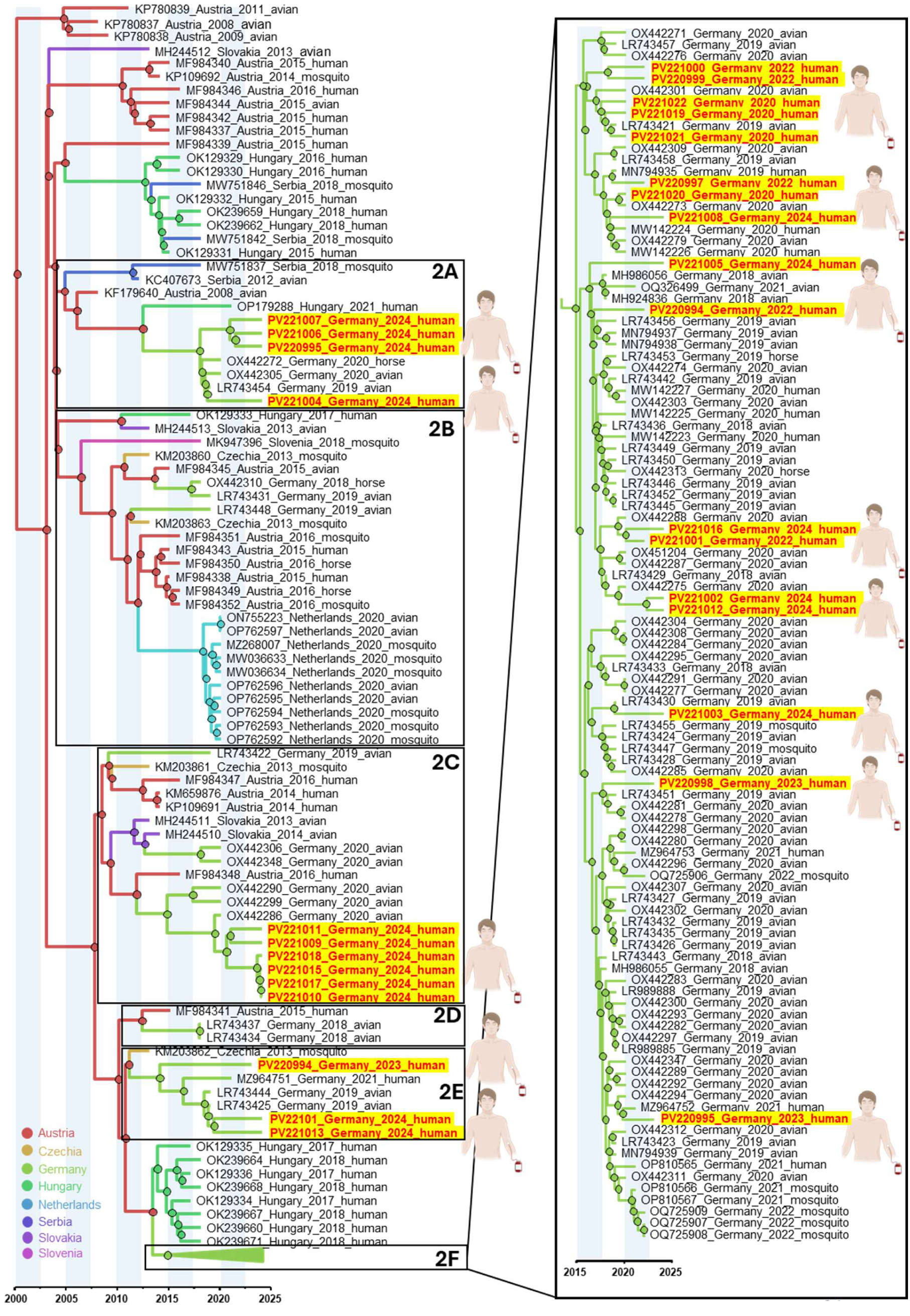
Bayesian maximum clade credibility (MCC) tree showing the time-scaled phylogeny of West Nile virus. This tree is based on complete or near-complete (≥70%) genome sequences, including the German sequences (subclades 2A–2F). The MCC tree was inferred using BEAST v1.10.5 with an uncorrelated relaxed molecular clock and a Skygrid coalescent tree prior, with model selection guided by stepping-stone sampling. Markov chain Monte Carlo (MCMC) chains were run for 200 million steps, sampling every 10,000 steps. Branch colors indicate the most probable geographic origin of descendant nodes (see color code). Subclades containing German WNV strains are displayed to the right of the tree. The German WNV strains from blood donors sequenced in this study are highlighted. The time scale is shown along the x-axis and represents years before the last sampling date (2024).

### Evolutionary dynamics and phylogeographic reconstruction of the WNV spread pattern in Germany

To investigate the emergence, introduction routes, and dispersal dynamics of West Nile virus in Germany, we applied both discrete-trait and continuous phylogeographic models. This integrative framework enabled reconstruction of WNV evolutionary history, identification of geographic transmission hubs, and quantification of viral migration patterns. Time-scaled Bayesian phylogenies placed the most recent common ancestor (TMRCA) of the German WNV-associated subclades around June 2015 (95% HPD: July 2014– March 2016), suggesting that the viruses seeding Germany shared a common ancestor with strains circulating in Hungary and Austria (Figure S8). Discrete and continuous phylogeographic analysis and Evolaps reconstruction (Figure 4) indicate that Germany did not experience a single introduction followed by in-country diversification, but rather multiple independent introductions of closely related viruses from Central European source populations (Figure 4-6; Figure S9).

**Figure 4.**
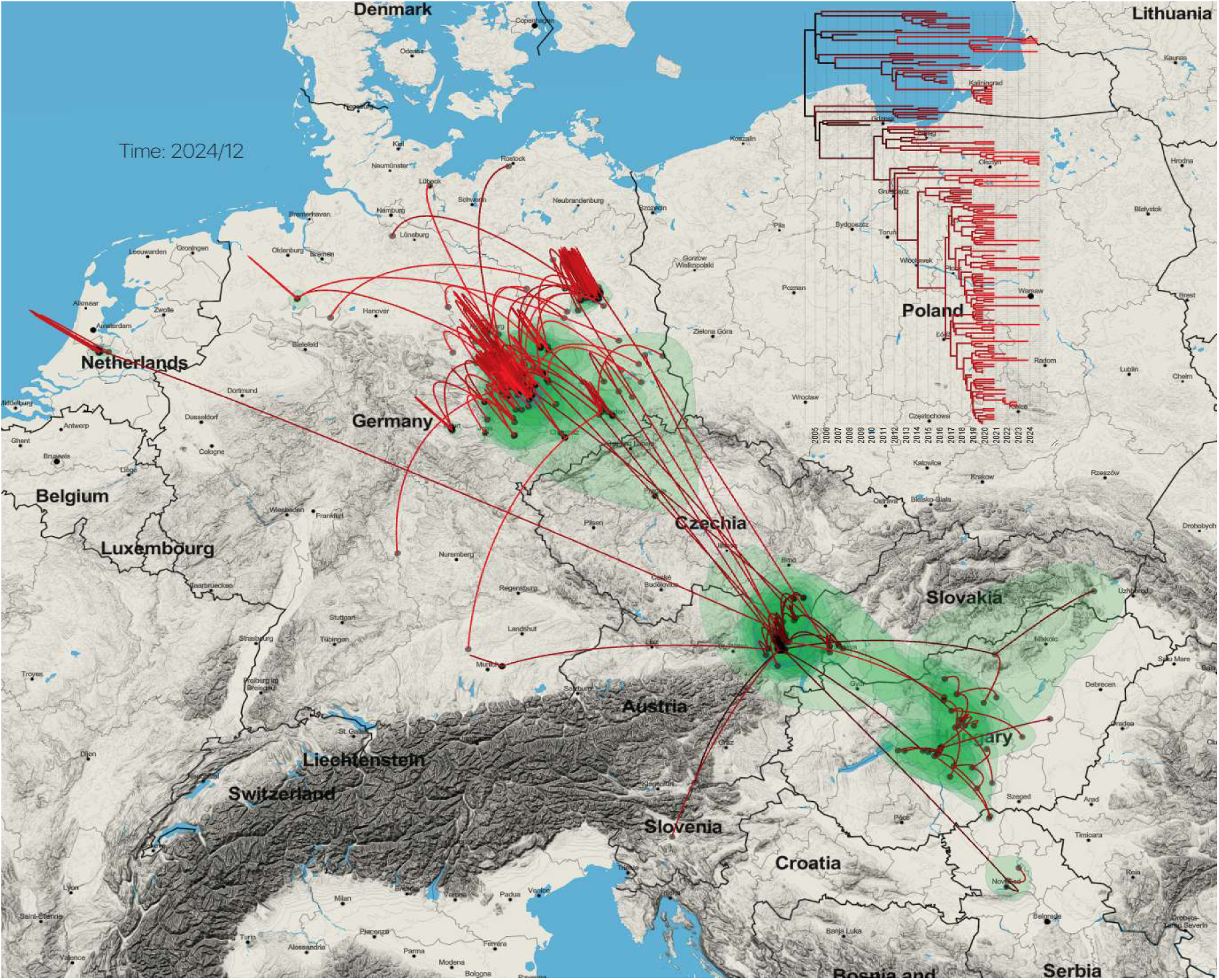
The phylogeographic reconstruction of West Nile virus origin and dispersal in Germany. The time-scaled maximum clade credibility (MCC) tree (top right) is based on ≥70% genome sequences (n = 170) of the subclades 2A-F. The phylogeny was inferred using a continuous Bayesian phylogeographic model based on 2,000 posterior trees. Viral dispersal is depicted on a geographic map, with branches representing inferred migration pathways and superimposed over the 80% highest posterior density (HPD) regions that reflect phylogeographic uncertainty. Branch colours indicate the time scale, ranging from black (time to the most recent common ancestor, TMRCA) to red (most recent sampling time). Green shaded areas represent the HPD regions, highlighting zones of concentrated viral activity and inferred transmission hubs. The inset panel (top left) displays the inferred geographic spread of WNV-2a as of December 2024.

This interpretation is supported by the existence of six distinct WNV subclades (2A–2F), which consist of German strains, each with its introduction timing and geographic dispersal pattern (Figure 5). Subclade 2A was probably introduced around late August 2018 from Austria. It has established sustained transmission in Brandenburg, Lower Saxony, Schleswig-Holstein, and Saxony-Anhalt, with a total diffusion range of ∼185 km and an estimated migration rate of ∼25.7 km/year for the whole subclade (Figure 5-6). Subclade 2B, introduced around September 2017, was detected only in 2018 and 20199 and has not been reidentified, suggesting failure to establish sustained local transmission. Based on our models, subclade 2C appeared in September 2015, with persistent detections in Berlin, Brandenburg, Thuringia, and Saxony since 2019. The subclade exhibits a moderate diffusion (∼240 km) and a migration rate of ∼36.9 km/year, consistent with stable local expansion. Subclade 2D, introduced probably around July 2018 in Bavaria, remained limited in distribution and lacked evidence of establishment or geographic spread beyond its origin. Subclade 2E, with an estimated introduction date near July 2016, has remained geographically restricted to Saxony-Anhalt and Brandenburg, with a modest spatial range (∼110 km) and migration rate of ∼18.9 km/year, suggesting localized persistence. Subclade 2F is the most geographically widespread and genetically diverse German WNV group, introduced in July 2016 and now dominant across Saxony, Saxony-Anhalt, Thuringia, and likely imported cases in Bavaria and North Rhine-Westphalia. This subclade has diffused over ∼380 km and shows the highest migration rate at ∼62.3 km/year. (Figure 5-6). Combined data from continuous diffusion models and k-means clustering (Figures 4–6) indicate that Brandenburg, Berlin, Saxony, and Saxony-Anhalt have acted as key transmission hubs supporting the local establishment and inter-regional spread of WNV. Brandenburg emerged as a central node, participating in multiple subclades (2A, 2C, 2E), and serving both as a recipient and source of virus dispersal to surrounding regions. Notably, Berlin, particularly in subclade 2C, has shown strong phylogenetic clustering with sequences from Brandenburg and Thuringia, suggesting its role as an epidemiological bridge between eastern and central Germany. The repeated detection of genetically related virus strains in Berlin across multiple seasons, along with its central role in transmission trajectories, supports its function as an urban maintenance focus. Similarly, Saxony and Saxony-Anhalt, especially within subclades 2F and 2E, have seeded westward spread toward Thuringia, where regional diversification and establishment occurred. In contrast, federal states such as Schleswig-Holstein and Lower Saxony showed sporadic detections with no evidence of sustained circulation or onward spread yet.

**Figure 5.**
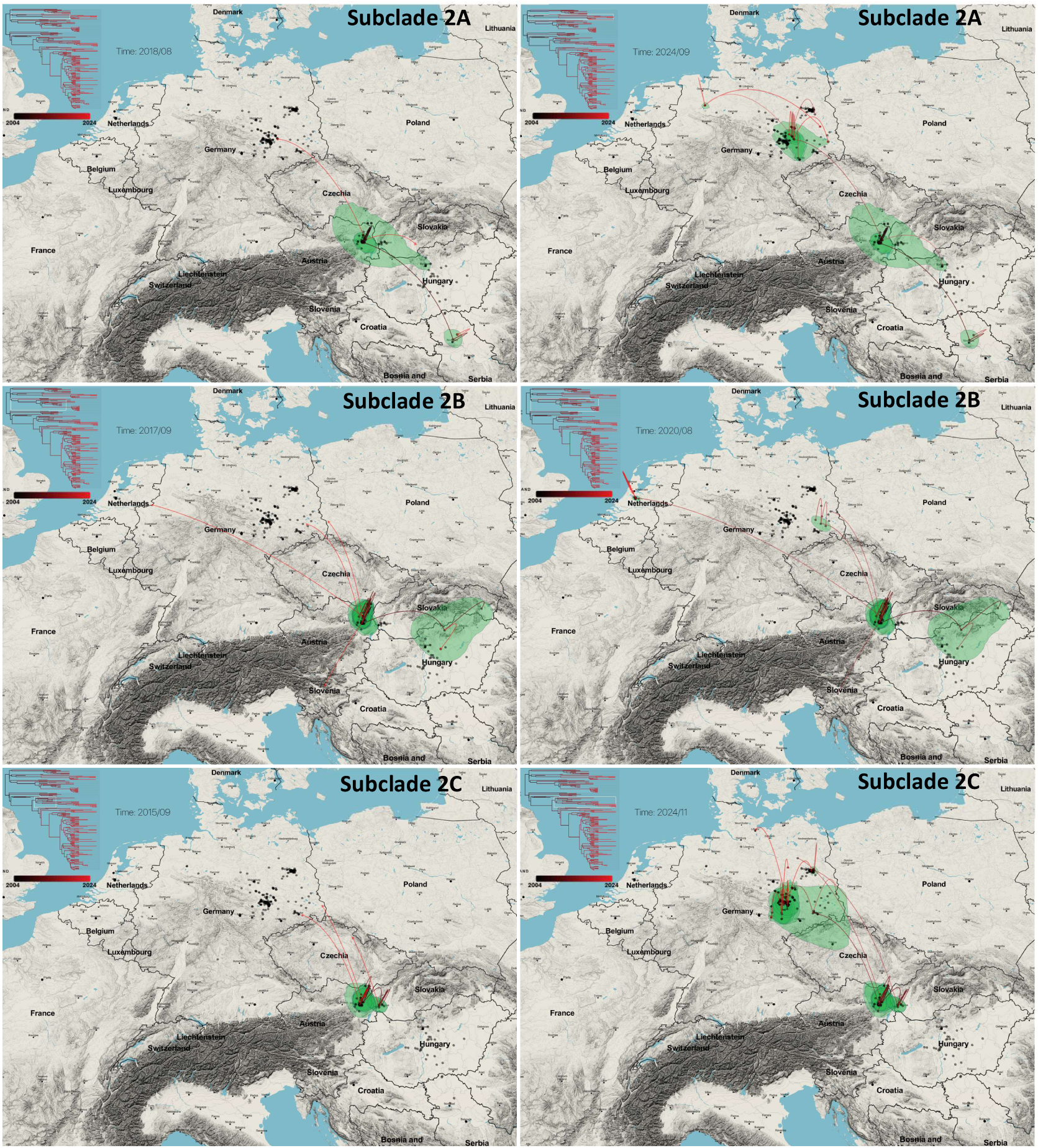
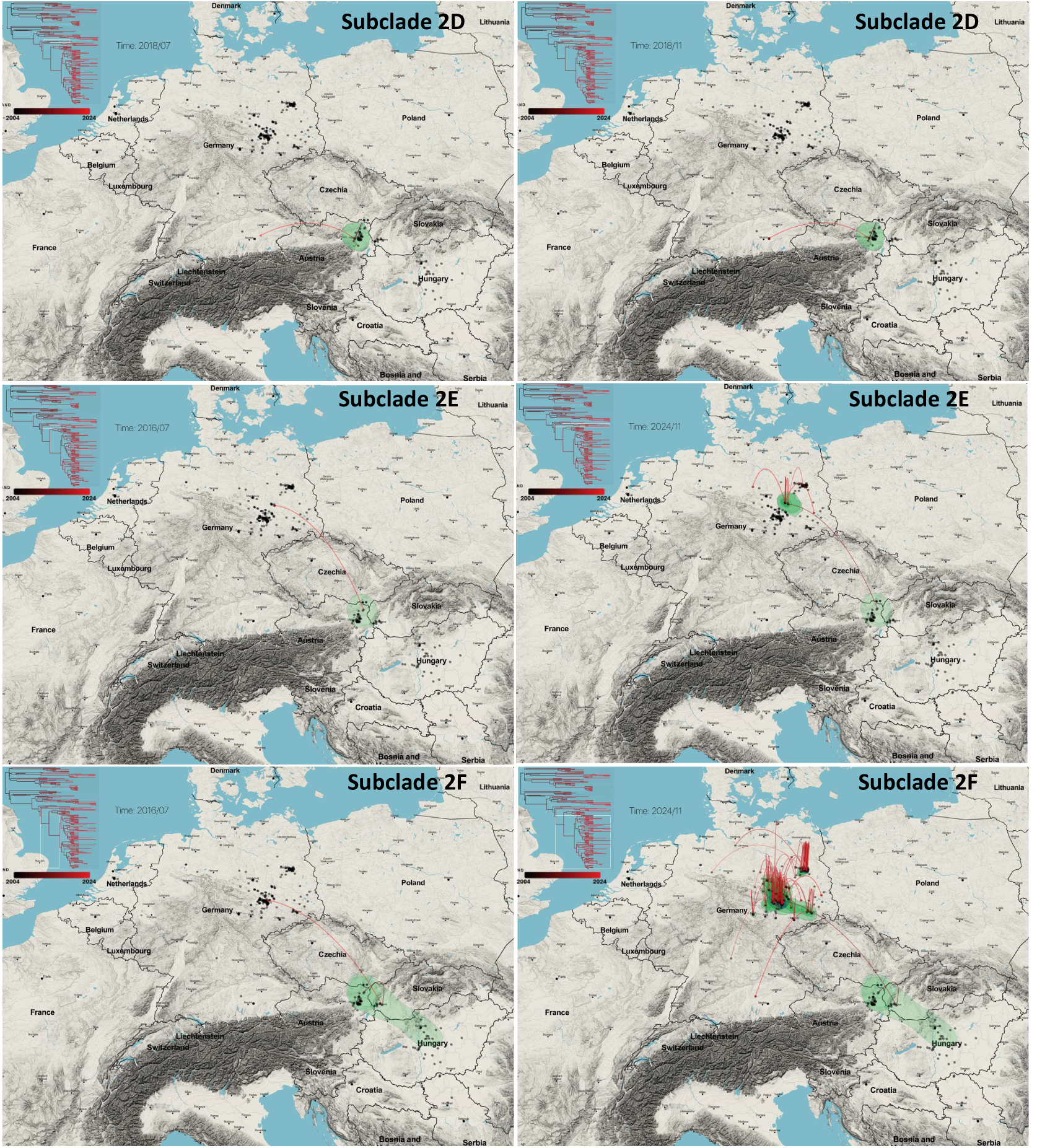
Spatial reconstruction of WNV subclades 2A–F based on continuous phylogeographic analysis. The time-scaled maximum clade credibility (MCC) tree (top left) is based on ≥70% genome sequences. Each dot represents a node from the maximum clade credibility (MCC) tree, while curved lines represent phylogenetic branches and illustrate the directionality of viral movement across geographic space. The left panel shows the estimated time of introduction (year/month) into Germany, while the right panels display the geographic distribution of each subclade.

**Figure 6.**
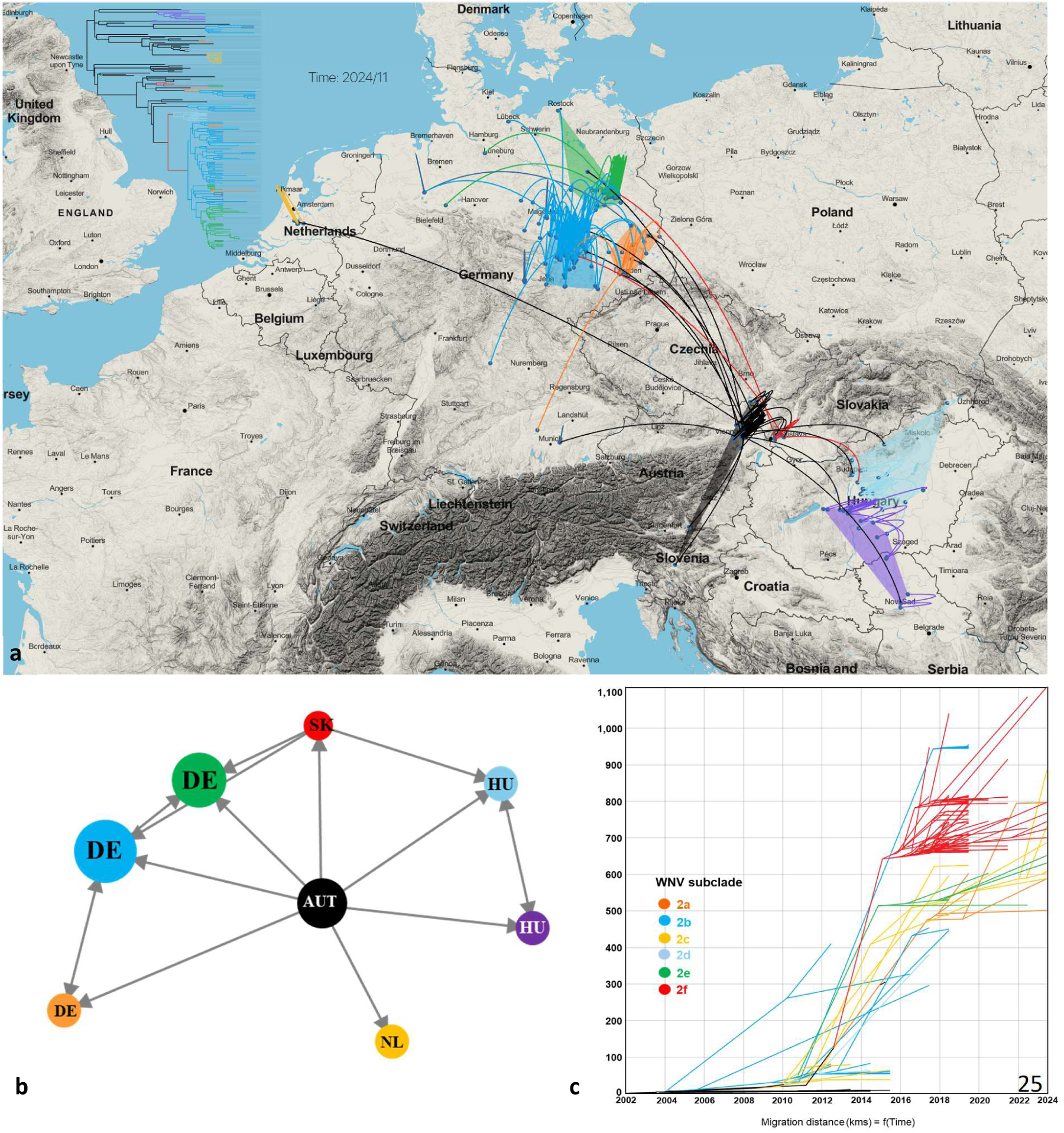
Spatial clustering analysis of WNV subclades 2A–F in Germany. (a) Locations associated with each node of the MCC tree are subjected to K-means clustering (k = 9) to define geographic zones and improve the visualization of the phylogeographic scenario. Localities are coloured according to their assigned cluster, and paths are coloured based on the cluster of origin. (b) The transition graph displays source-to-destination relationships between and within clusters, with nodes labelled by country names. Node size is proportional to the number of localities in each cluster. (c) The migration distance curve shows the distribution of geographic distances associated with dispersal events, highlighting the relative contributions of local versus long-distance spread in shaping the population structure of West Nile virus in Germany.

## Discussion

The diagnostic and epidemiological challenges associated with WNV infections, particularly among asymptomatic patients, require sensitive and scalable genome sequencing methods that can be integrated into routine public health surveillance and transfusion safety systems. These challenges are particularly pronounced in blood donor surveillance, where viremia is typically low, often resulting in suboptimal genome recovery for molecular epidemiological analyses [9, 20]. To address these limitations, we developed and validated a highly sensitive amplicon-based next-generation sequencing (NGS) protocol, optimized for low RNA input and high Ct value samples. This approach refines previously developed amplicon-based protocols for WNV genome recovery and other orthoflaviviruses [12–18], and our results show it performs well even with blood donor samples carrying Ct values up to 35 (less than 10 copies/µL). However, we observed that successful genomic recovery from blood donor samples, as well as clinical specimens, is strongly influenced by the overall quality of the sample material. Even among samples with similar Ct values, differences in storage conditions, RNA integrity, and sample handling impacted sequencing success. This reinforces the need for standardized sample processing protocols when applying genomic tools in routine surveillance or diagnostic workflows. Although we used the smallest Illumina sequencing platform (iSeq100) for our experiments, the protocol is compatible with any sequencing platform that supports 300-cycle runs (2 × 150 bp), as the designed amplicon lengths fall well within this read range. Using this method, we sequenced a large panel of WNV genomes collected from blood donors between 2020 and 2024, revealing six distinct subclades circulating in Germany (2A–2F). These represent separate introduction events, primarily originating from Central Europe, especially Austria and Slovakia, as supported by discrete trait phylogeography and continuous diffusion models. Several of these subclades, including 2F and 2C, have become established and are undergoing regional diversification, particularly in Brandenburg, Berlin, Saxony, and Saxony-Anhalt. These regions now act as long-term transmission hubs, maintaining local foci and facilitating onward spread to the northwest. Berlin emerged as a particularly important urban transmission node with reported focal establishment of WNV [29], showing repeated seasonal detections and strong phylogenetic linkage to nearby federal states. Our findings include the detection of WNV-positive blood donor samples in federal states not previously considered endemic, such as North Rhine-Westphalia, Bavaria, and Schleswig-Holstein. While these observations could represent true introduction and transmission events, the lack of systematic documentation of nation travel histories among blood donors complicates the differentiation between locally acquired and imported infections. In three of these cases, two from Bavaria and one from North Rhine-Westphalia, a recent stay in WNV-endemic regions was documented, supporting the hypothesis of infection through national travel-related introduction rather than local transmission. A standardized approach to collecting all travel histories during the follow-up of WNV-positive blood donors would substantially improve the resolution of future surveillance and epidemiological analyses. Phylogeographic reconstructions indicate that multiple WNV lineage 2 subclades were introduced into Germany as early as 2016–2018, predating previous estimates. This suggests that undetected transmission occurred before the first confirmed autochthonous cases, despite efforts to identify hidden WNV infections in at-risk populations [30]. This highlights the value of retrospective genomic analysis for tracing virus emergence. The presence of a unique mutation in the helicase domain of the NS3 protein in subclade 2F, which now dominates the German WNV landscape, suggests possible adaptive changes relevant to viral fitness, though functional studies are needed to confirm this. Our study also reinforces the role of blood donors as effective sentinels for monitoring WNV activity. Donors are systematically screened and provide geographically widespread, temporally dense sampling across the country. Prior studies have shown that blood donor screening can detect WNV circulation ahead of symptomatic cases, providing a sensitive early-warning signal for public health response [19,20]. Our findings extend this value by demonstrating that donor-derived samples can also yield high-quality genomic data, enabling phylogenetic and phylogeographic inference, even with low viral load. Genomic data from blood donors thus serve not only diagnostic or confirmatory purposes, but also power surveillance systems to understand WNV introduction routes, local evolution, and the impact of environmental drivers such as climate and vector ecology [31]. This is especially relevant considering the predicted northward expansion of mosquito habitats under climate change, which has already been associated with WNV spread into previously unaffected regions of Europe [32]. Our findings are timely given the accelerating impact of climate change on the spread and intensity of mosquito-borne diseases in Europe [31]. Warmer temperatures and extended vector seasons are expanding the ecological niche of WNV and other arboviruses, necessitating proactive molecular surveillance to detect early incursions and inform vector control interventions. This study provides a blueprint for future genomic surveillance of WNV in Germany and beyond. It demonstrates that even in settings with low virus titres and decentralized testing structures, whole-genome sequencing is feasible and epidemiologically valuable when supported by sensitive and flexible molecular protocols. The validated sequencing protocol and bioinformatic workflows are publicly available and can be adopted or adapted by other laboratories in the EU for harmonized surveillance efforts.

## Conclusion

This study demonstrates that integrating amplicon-based sequencing with blood donor screening enables early, accurate, and geographically extensive genomic surveillance of WNV. This combined approach strengthens outbreak preparedness, supports timely detection of viral introductions, informs risk-based public health responses, and advances our understanding of WNV evolutionary dynamics across Europe.

## Data Availability

The genomic sequences obtained in this study are available in the GenBank under accession numbers PV220993-PV221022, and the corresponding raw sequencing data were deposited in the Sequence Read Archive (SRA) under Bioproject ID PRJNA1223181.

https://www.protocols.io/view/west-nile-virus-orthoflavivirus-nilense-lineage-2-q26g71q98gwz/v1

## Ethical statement

This study was conducted in accordance with the ethical principles outlined in the Declaration of Helsinki (revised 2013). All blood donor samples, and associated metadata used in this study were fully anonymized prior to analysis. The legal basis for the collection and analysis of these samples is established by Section 28 of the German Medicinal Products Act (Arzneimittelgesetz, AMG), Section 14 of the Transfusion Act (Transfusionsgesetz, TFG), and Sections 4 and 7 of the Infection Protection Act (Infektionsschutzgesetz, IfSG). Section 7 IfSG mandates the reporting of WNV infections, while Section 4 IfSG provides the framework for the analysis of such data. Case reporting and laboratory-based surveillance are further governed by Sections 63i AMG and 19 TFG. As these activities are part of official public health responsibilities under German national legislation, no separate ethics committee approval was required, and no institutional ethics review was sought.

## Funding statement

The mNGS investigations were funded by the German Federal Ministry of Education and Research (BMBF) within the project PREPMedVet no. 13N15449 whereas the amplicon-based sequencing approach within Culifo3 project no. 2819107A22 by the German Federal Ministry of Food and Agriculture.

## Data availability

The genomic sequences obtained in this study are available in the GenBank under accession numbers PV220993–PV221022, and the corresponding raw sequencing data were deposited in the Sequence Read Archive (SRA) under Bioproject ID PRJNA1223181.

## Conflict of interest statement

None.

## Acknowledgements

The authors would like to thank all participating blood establishments for reporting cases and providing samples of WNV-positive blood donations. We also acknowledge the regional public health authorities for their collaboration in case investigation, data provision, and support of surveillance activities during the West Nile virus transmission seasons in Germany. We are grateful to Unchana Lange for her excellent technical assistance

## Authors’ contributions

GET, JS-C, and DC: conceptualization; GET, MS, RO, RL, MS, DC: data collection; GET, MP, MS, AB, HB: laboratory analysis; GET, MS, AT, BH, RL, DC: data analysis; GET, JS-C, and DC: writing and editing; GET, MS, AT, BH, PS, AH, SJ, RO, RL, MS, JS-C, DC: manuscript review.

## Notes

### Competing Interest Statement

The authors have declared no competing interest.

