## Supplementary material for "Blood donors as sentinels for genomic surveillance of West Nile virus in Germany (2020–2024) using a sensitive amplicon-based sequencing approach"

**Supplementary Figure S1**. Pooling strategy for West Nile virus genome sequencing primers and the distribution of amplicon numbers by fragment length.

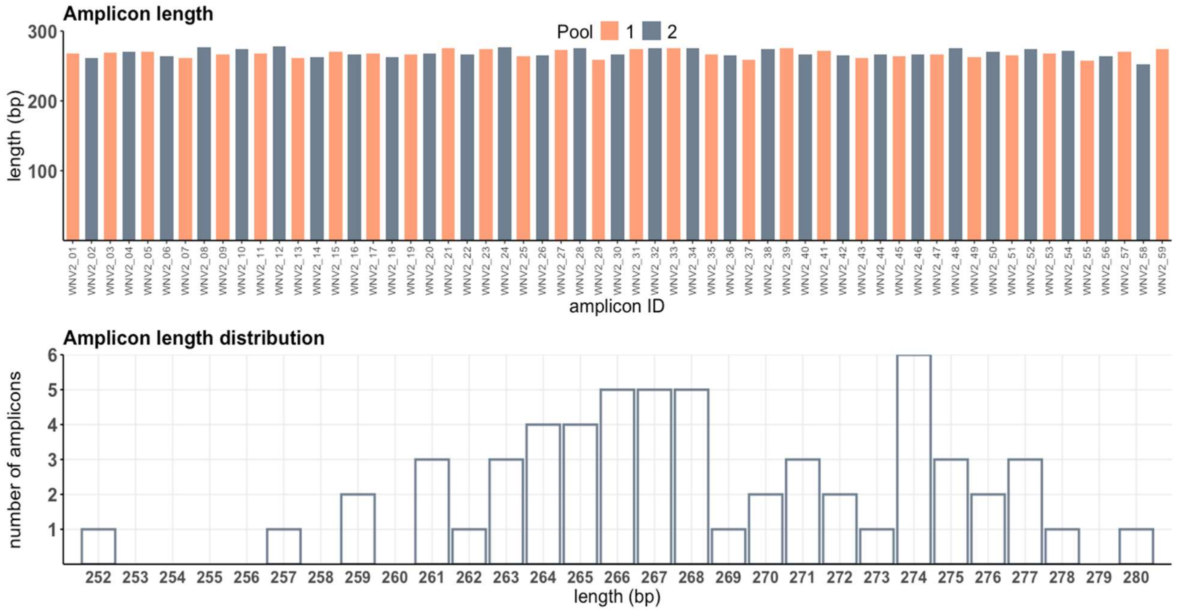

**Supplementary Figure S2**. Schematic representation of the proposed workflow for West Nile virus amplicon-based sequencing. The workflow outlines each step from sample processing to results, including both wet lab procedures and bioinformatic analyses.

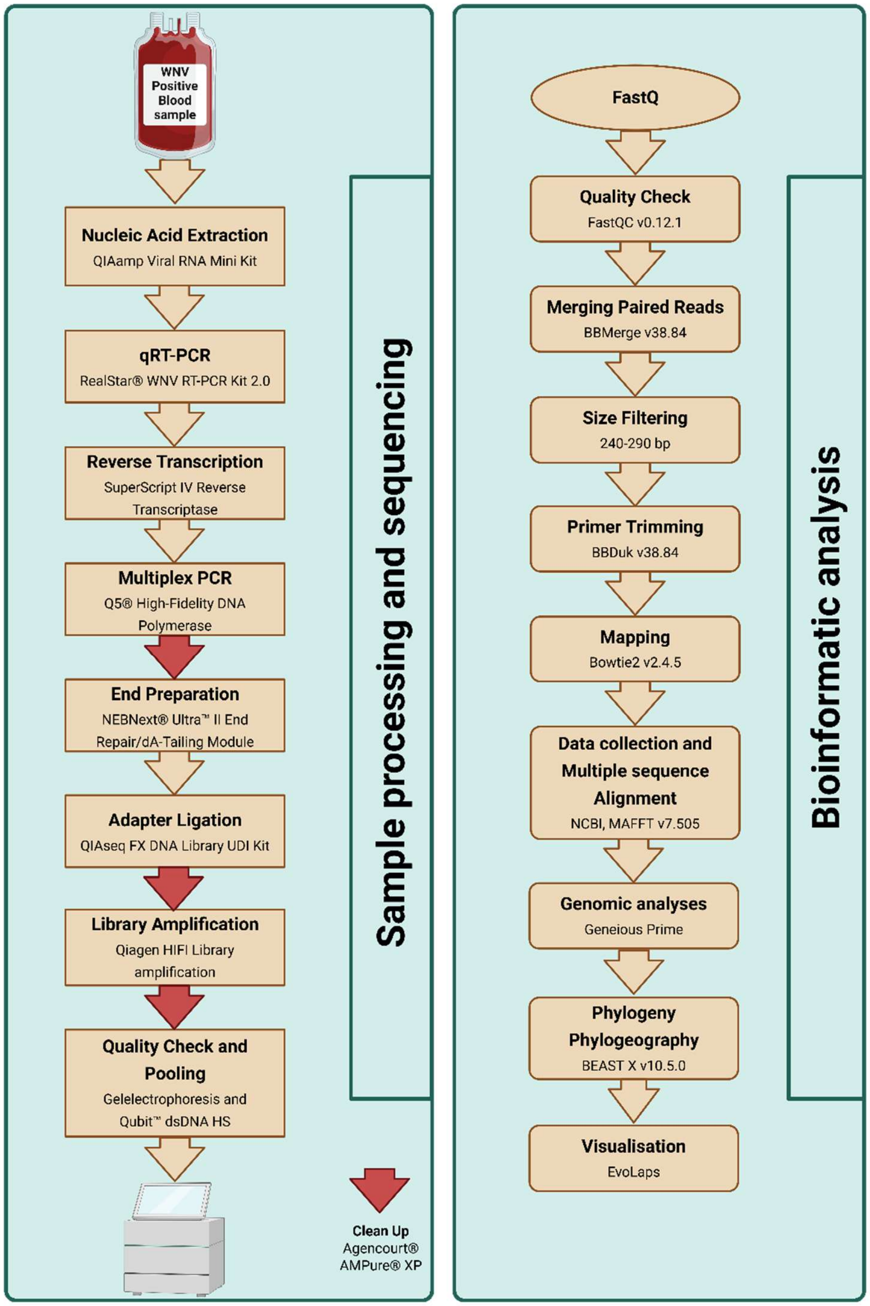

**Supplementary Figure S3**. Sequencing performance metrics obtained from West Nile virus isolates (BNI-129; B956; UG37) during the optimization of the amplicon-seq approach for WNV genome sequencing: **a)** Ct values and estimated viral copy numbers (copies/µl) from serial dilutions; **b)** sequencing results including read counts per serial dilution; **c)** genome recovery percentages at 10x coverage; **d)** merged/non-merged reads, discarded reads, clean reads, mapped and unmapped reads related to each dilution of WNV reference strains.

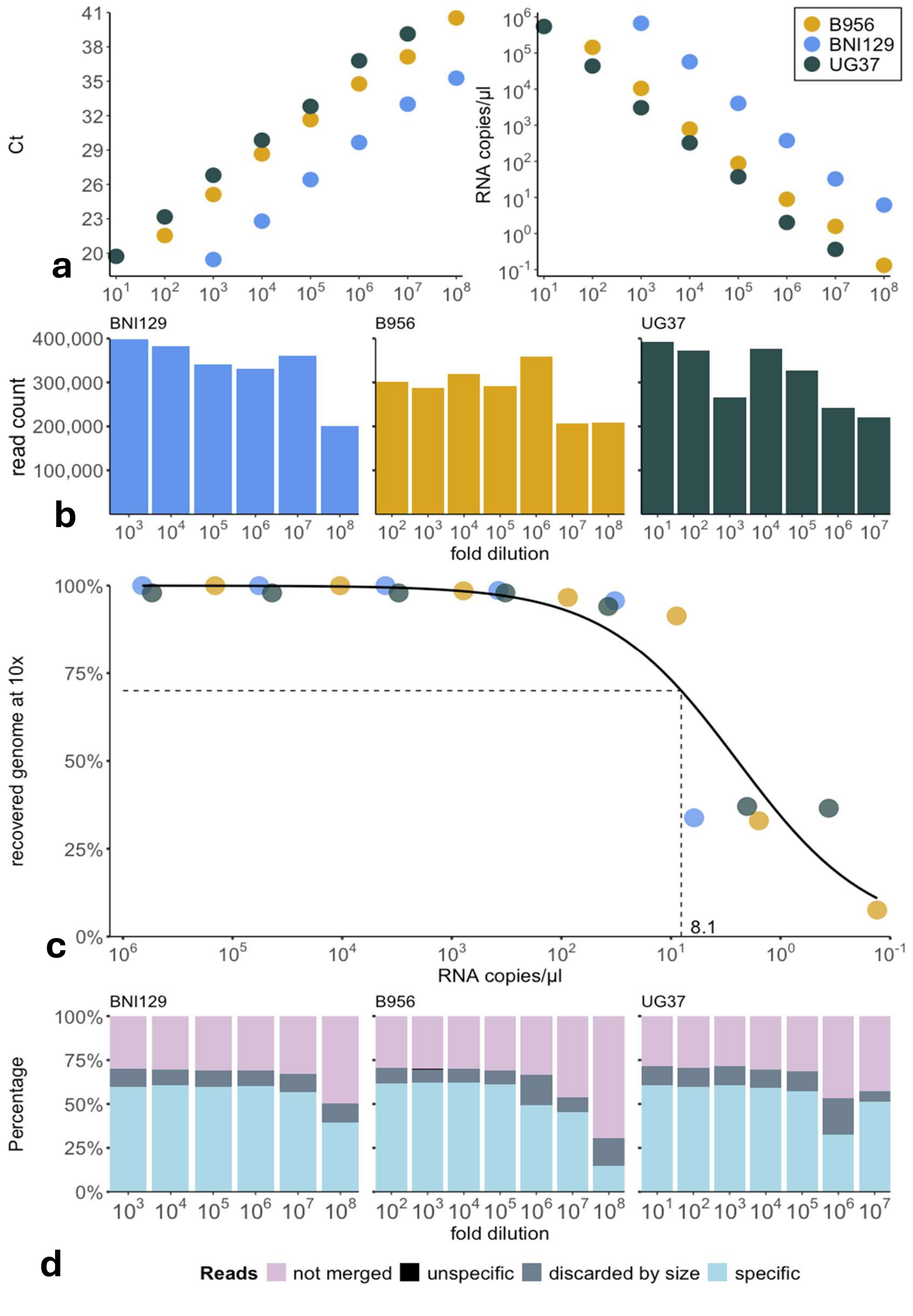

**Supplementary Figure S4**. Sequencing performance metrics obtained from blood donor samples during the optimization of the amplicon-seq approach for WNV genome sequencing. **a)** sequencing results including read counts for each sample; **b)** merged/non-merged reads, discarded reads, clean reads, mapped and unmapped reads related to each WNV positive blood donor sample according to their Ct values/copies/µL; **c)** The viral RNA concentration threshold required for achieving ≥70% genome recovery was estimated using a generalized linear model (GLM) with a logit link function and a quasibinomial error distribution to account for overdispersion. A genome completeness cutoff of 70% was selected based on prior *Orthoflavivirus* studies as a practical threshold for enabling reliable phylogeographic inference.

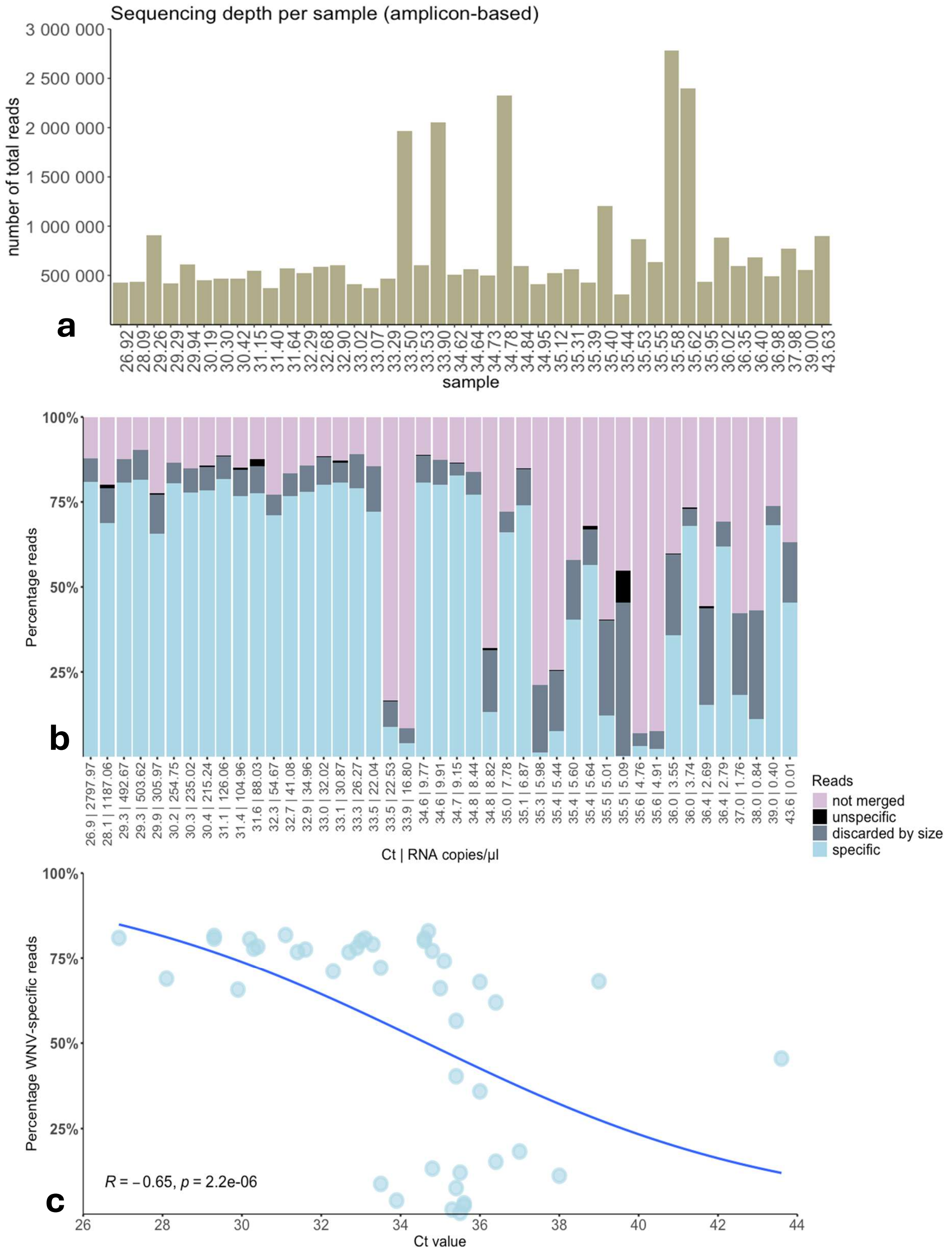

**Supplementary Figure S5.** Summarized breadth of coverage across the WNV genome for positive blood donor samples sequenced using the amplicon-based approach. The *x*-axis indicates genome positions, and the *y*-axis shows the total read depth for each sample. A schematic representation of the WNV genome is included, showing the amplified regions corresponding to each primer pair used for genomic coverage.

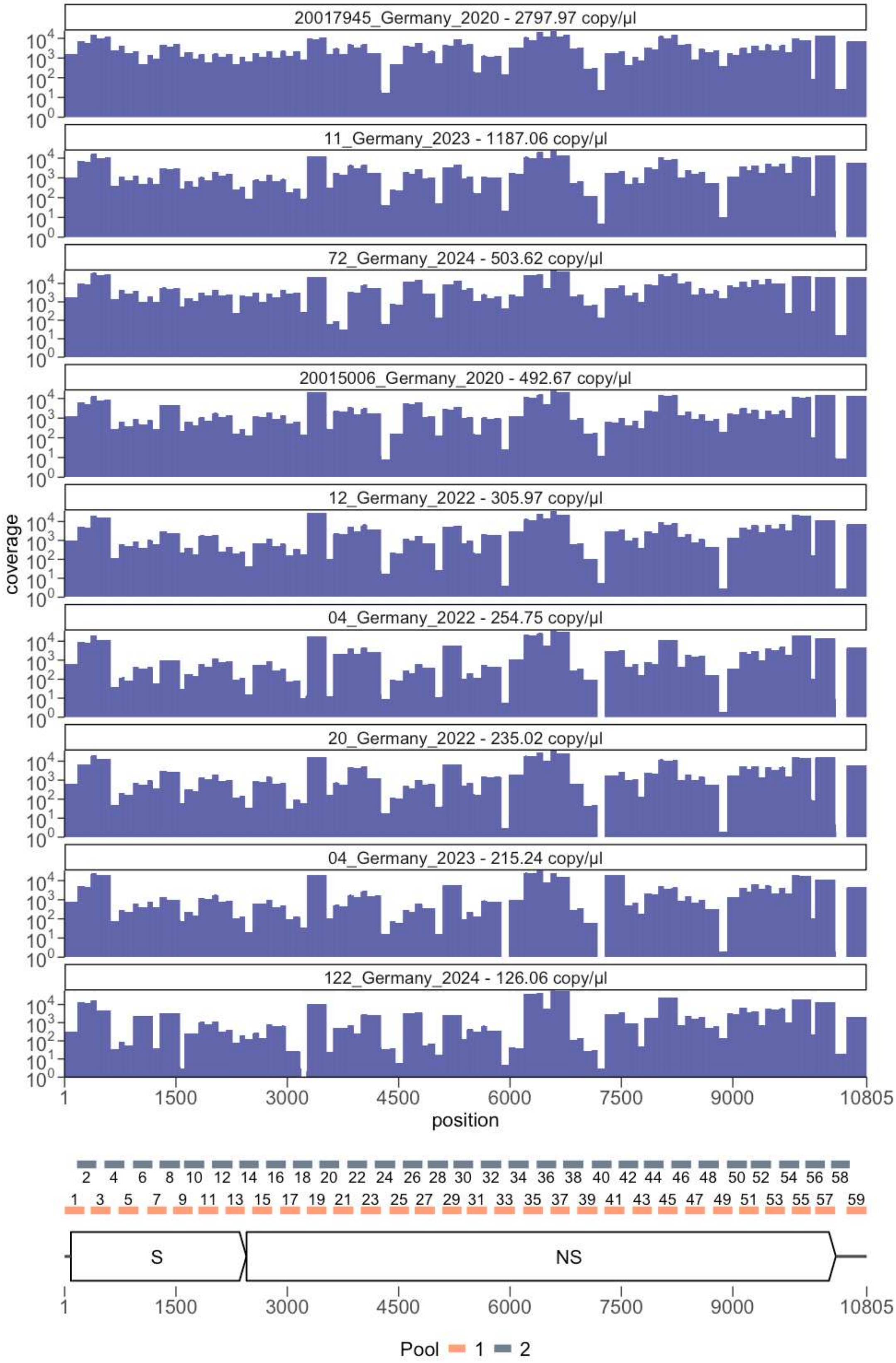
20017945_Germany_2020 - 2797.97 copy/µl
11_Germany_2023 - 1187.06 copy/µl
72_Germany_2024 - 503.62 copy/µl
20015006_Germany_2020 - 492.67 copy/µl
12_Germany_2022 - 305.97 copy/µl
04_Germany_2022 - 254.75 copy/µl
20_Germany_2022 - 235.02 copy/µl
04_Germany_2023 - 215.24 copy/µl
122_Germany_2024 - 126.06 copy/µl
coverage
position
S
NS
Pool 1 2

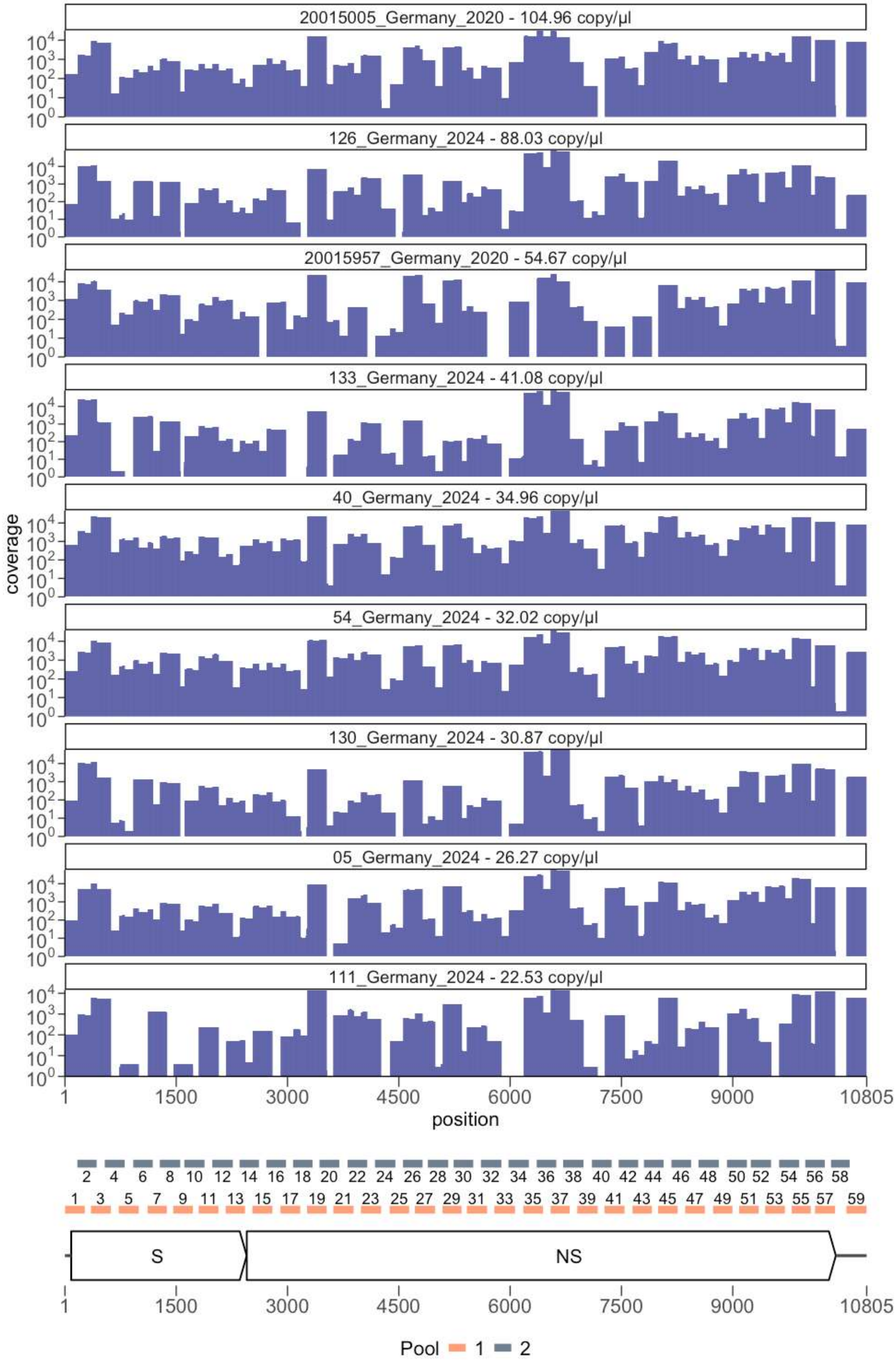

20015005_Germany_2020 - 104.96 copy/µl
126_Germany_2024 - 88.03 copy/µl
20015957_Germany_2020 - 54.67 copy/µl
133_Germany_2024 - 41.08 copy/µl
40_Germany_2024 - 34.96 copy/µl
54_Germany_2024 - 32.02 copy/µl
130_Germany_2024 - 30.87 copy/µl
05_Germany_2024 - 26.27 copy/µl
111_Germany_2024 - 22.53 copy/µl
coverage
position
1 1500 3000 4500 6000 7500 9000 10805
S
NS
Pool 1 2

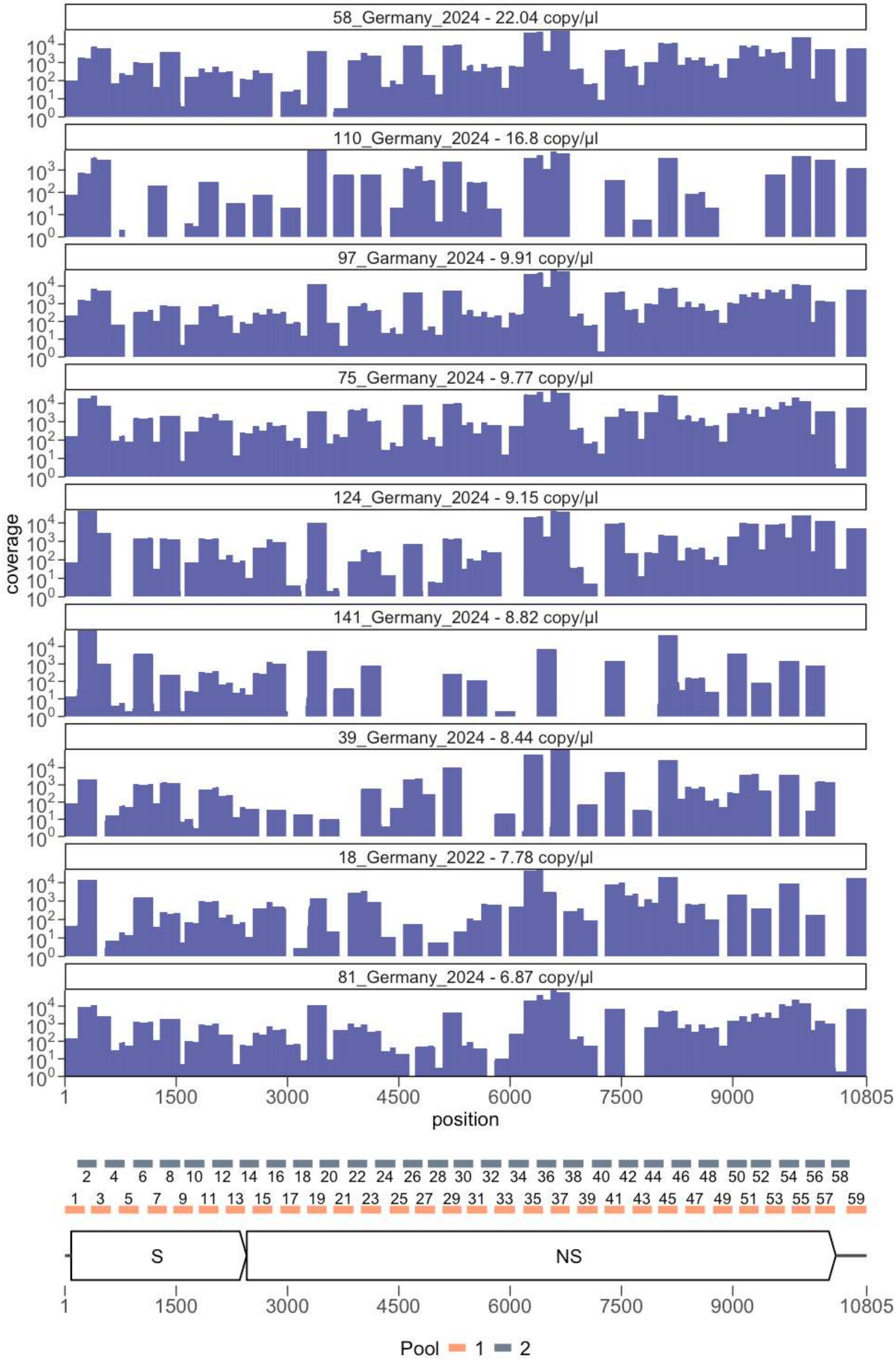
58_Germany_2024 - 22.04 copy/µl
110_Germany_2024 - 16.8 copy/µl
97_Garmany_2024 - 9.91 copy/µl
75_Germany_2024 - 9.77 copy/µl
124_Germany_2024 - 9.15 copy/µl
141_Germany_2024 - 8.82 copy/µl
39_Germany_2024 - 8.44 copy/µl
18_Germany_2022 - 7.78 copy/µl
81_Germany_2024 - 6.87 copy/µl
coverage
position
S
NS
Pool 1 2

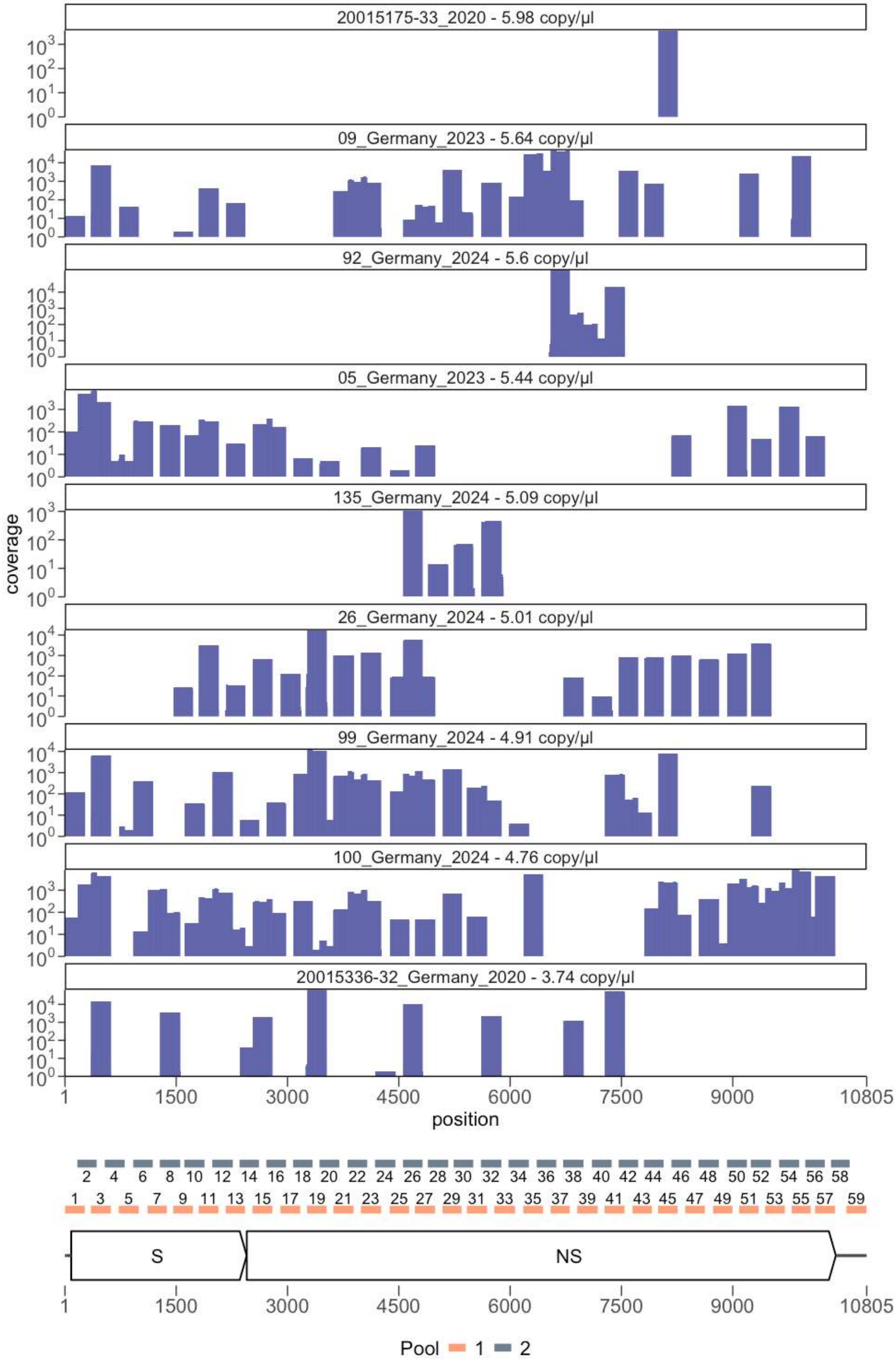
20015175-33_2020 - 5.98 copy/µl
09_Germany_2023 - 5.64 copy/µl
92_Germany_2024 - 5.6 copy/µl
05_Germany_2023 - 5.44 copy/µl
135_Germany_2024 - 5.09 copy/µl
26_Germany_2024 - 5.01 copy/µl
99_Germany_2024 - 4.91 copy/µl
100_Germany_2024 - 4.76 copy/µl
20015336-32_Germany_2020 - 3.74 copy/µl
coverage
position
1
1500
3000
4500
6000
7500
9000
10805
S
NS
Pool 1 2

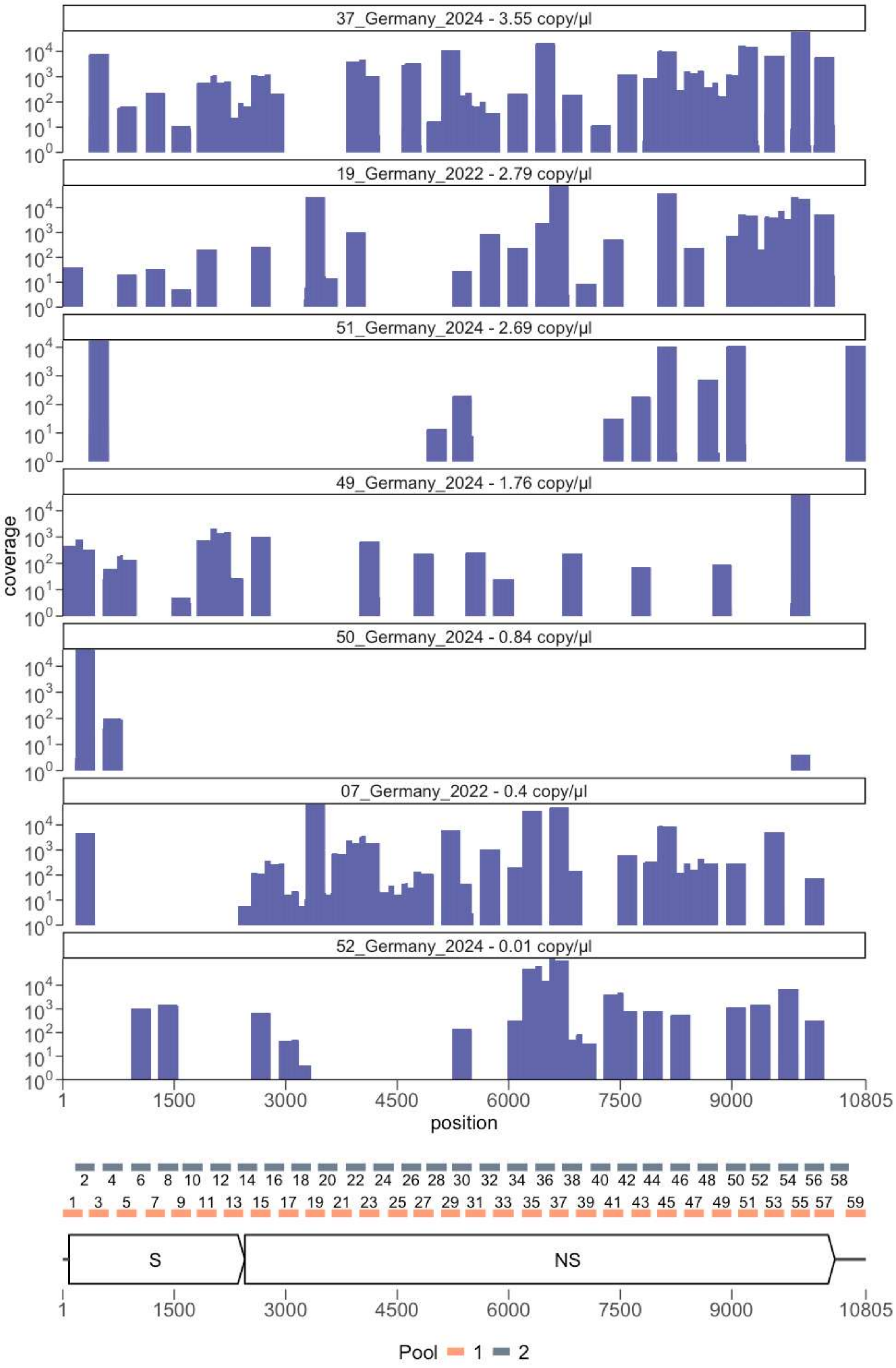

37_Germany_2024 - 3.55 copy/µl
19_Germany_2022 - 2.79 copy/µl
51_Germany_2024 - 2.69 copy/µl
49_Germany_2024 - 1.76 copy/µl
50_Germany_2024 - 0.84 copy/µl
07_Germany_2022 - 0.4 copy/µl
52_Germany_2024 - 0.01 copy/µl
coverage
position
S
NS
Pool 1 2

**Supplementary Figure S6.** Comparison between targeted amplicons and metagenomic sequencing for WNV-positive blood donor samples. Panel a illustrates the sequencing depth achieved using both methods, while panel b compares percent genome coverage at 1x and 10x.

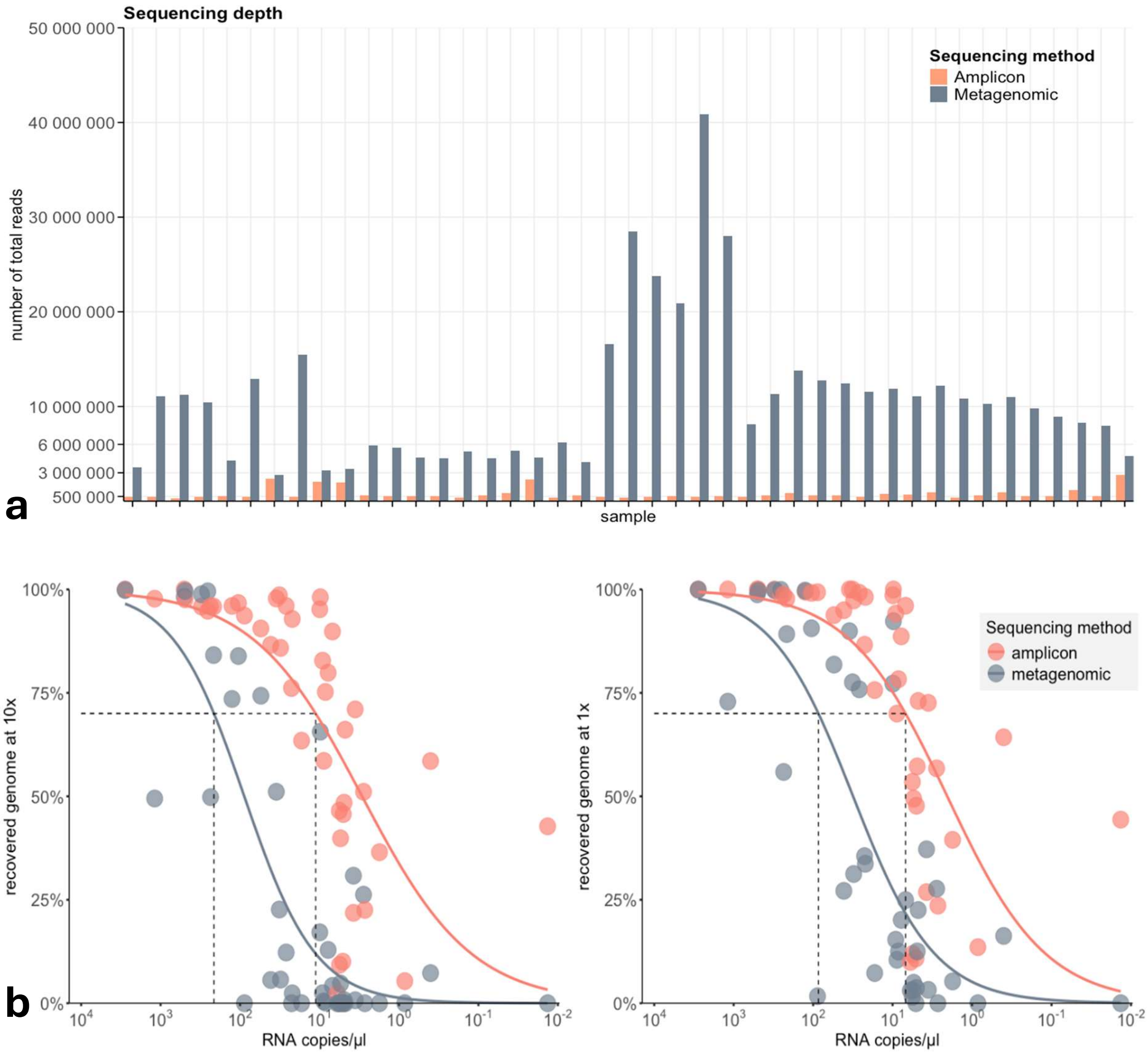

**Supplementary Figure S7.** Schematic representation of the West Nile virus genome, depicting characteristic amino acid substitutions of the subclades 2A-F. Amino acid changes are indicated by their position numbers and single-letter codes along the viral polyprotein, mapped to their respective genomic regions. Created in BioRender.

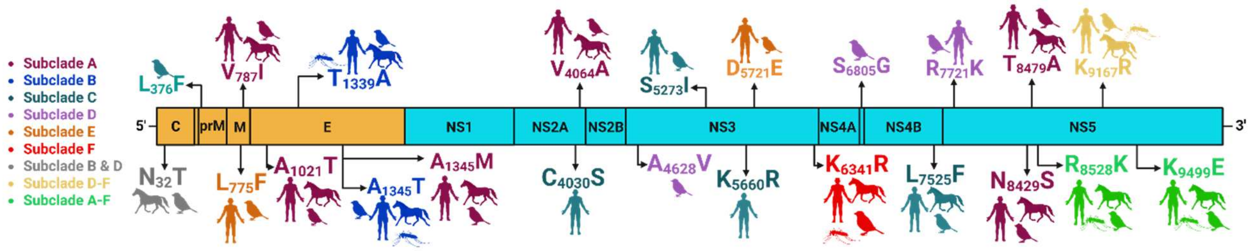

**Supplementary Figure S8.** Bayesian maximum clade credibility (MCC) tree showing the time-scaled phylogeny of West Nile virus lineage 2a in Europe. The tree is based on complete or near complete (≥70%) genome sequences. Branch colors indicate the most probable geographic origin of descendant nodes (see color code). Subclades containing German WNV strains are labeled to the right of the tree. The time scale is shown along the x-axis and represents years before the last sampling date (2024).

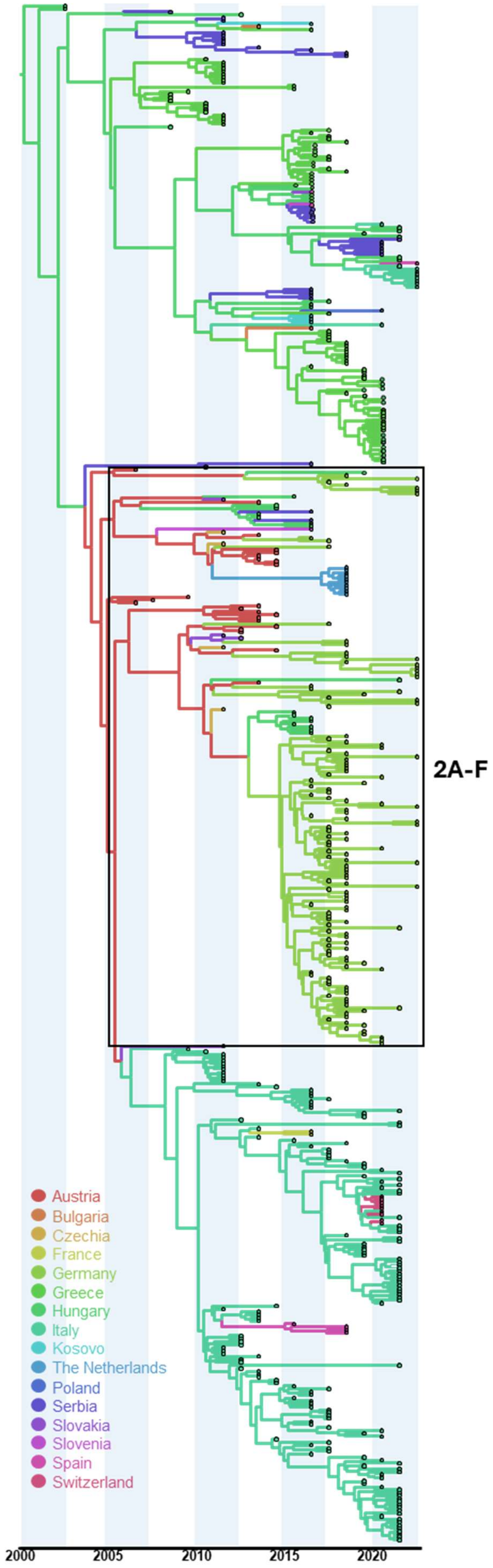
2A-F
Austria
Bulgaria
Czechia
France
Germany
Greece
Hungary
Italy
Kosovo
The Netherlands
Poland
Serbia
Slovakia
Slovenia
Spain
Switzerland
2000
2005
2010
2015
2020

**Supplementary Figure 8.** The phylogeographic reconstruction of West Nile virus origin and dispersal in Europe. The time-scaled maximum clade credibility (MCC) tree (left) is based ≥70% of genome sequences (n = 520) collected across Europe. The phylogeny was inferred using a continuous Bayesian phylogeographic model based on 4,000 posterior trees. Viral lineage dispersal is depicted on a geographic map, with branches representing inferred migration pathways and superimposed over the 80% highest posterior density (HPD) regions that reflect phylogeographic uncertainty. Branch colors indicate the time scale, ranging from black (time to the most recent common ancestor, TMRCA) to red (most recent sampling time). Green shaded areas represent the HPD regions, highlighting zones of concentrated viral activity and inferred transmission hubs. The inset panel displays the inferred geographic spread of WNV-2a as of December 2024.

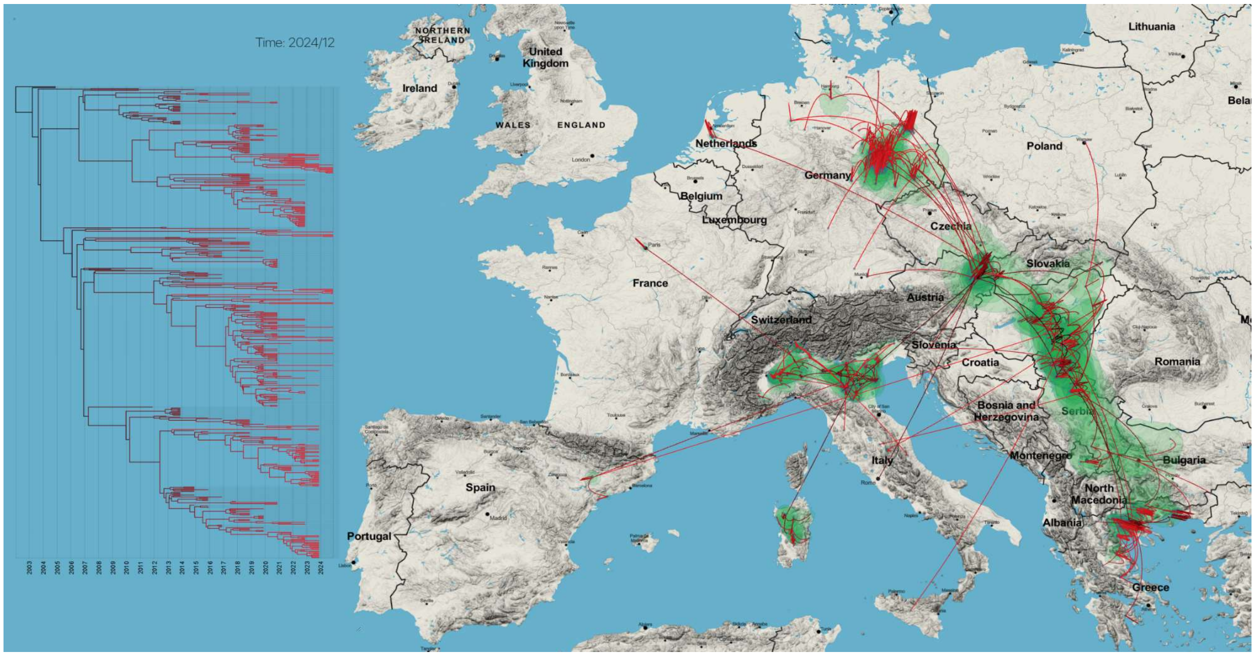

**Supplementary Table S1. Metadata and sequencing results for West Nile virus-positive blood donor samples from Germany (2020–2024), analyzed using the amplicon-seq approach.** The phylogenetic placement of these samples was determined and assigned to subclades within the West Nile virus lineage 2 phylogeny.

| Sample ID | Date of sampling | Federal State | Travel history | Genomic recovery ≥ 70% | Phylogenetic placement |
|---|---|---|---|---|---|
| 20015005 | Aug-2020 | Saxony | no | yes | Subclade 2F |
| 20015006 | Aug-2020 | Saxony | unknown | yes | Subclade 2F |
| 20015175-33 | Aug-2020 | Saxony | unknown | no | Subclade 2C |
| 20015336-32 | Aug-2020 | Berlin | Brandenburg | no | Subclade 2F |
| 20015957 | Sep-2020 | Saxony | no | yes | Subclade 2F |
| 20017945 | Sep-2020 | Saxony-Anhalt | no | yes | Subclade 2F |
| 2022-04 | Aug-2022 | Saxony-Anhalt | no | yes | Subclade 2E |
| 2022-07 | Aug-2022 | Saxony | no | yes | Subclade 2F |
| 2022-12 | Aug-2022 | Saxony-Anhalt | no | yes | Subclade 2F |
| 2022-18 | Aug-2022 | Bavaria | Endemic areas in Eastern Germany | yes | Subclade 2F |
| 2022-19 | Aug-2022 | Saxony-Anhalt | Croatia | no | Subclade 2F |
| 2022-20 | Aug-2022 | Saxony-Anhalt | Slovenia | yes | Subclade 2F |
| 2023-04 | Jul-2023 | Saxony Anhalt | no | yes | Subclade 2E |
| 2023-05 | Jul-2023 | North Rhine-Westphalia | Greece, Berlin | yes | Subclade 2F |
| 2023-09 | Jul-2023 | Thuringia | no | no | Subclade 2F |
| 2023-11 | Aug-2023 | Saxony-Anhalt | no | yes | Subclade 2F |
| 2024-05 | Jul-2024 | Saxony | no | yes | Subclade 2A |
| 2024-26 | Aug-2024 | Schleswig-Holstein | unknown | no | Subclade 2A |
| 2024-37 | Aug-2024 | Saxony | no | yes | Subclade 2F |
| 2024-39 | Aug-2024 | Bavaria | Slovakia, Italy | yes | Subclade 2F |
| 2024-40 | Aug-2024 | Saxony-Anhalt | Fuerteventura | yes | Subclade 2A |
| 2024-49 | Aug-2024 | Saxony-Anhalt | no | no | Subclade 2F |
| 2024-50 | Aug-2024 | North Rhine-Westphalia | Hamburg | no | Subclade 2F |
| 2024-51 | Aug-2024 | Lower Saxony | no | no | Subclade 2A |
| 2024-52 | Aug-2024 | Saxony-Anhalt | Brandenburg | no | Subclade 2F |
| 2024-54 | Aug-2024 | Saxony-Anhalt | no | yes | Subclade 2F |
| 2024-58 | Aug-2024 | Lower Saxony | no | yes | Subclade 2A |
| 2024-72 | Aug-2024 | Brandenburg | no | yes | Subclade 2A |
| 2024-75 | Aug-2024 | Brandenburg | no | yes | Subclade 2F |
| 2024-81 | Aug-2024 | Brandenburg | no | yes | Subclade 2C |
| 2024-92 | Aug-2024 | Schleswig-Holstein | no | no | Subclade 2A |
| 2024-97 | Aug-2024 | Saxony | no | yes | Subclade 2C |
| 2024-99 | Sep-2024 | Saxony | no | no | n/a |
| 2024-100 | Sep-2024 | Thuringia | no | yes | Subclade 2C |
| 2024-110 | Sep-2024 | Saxony-Anhalt | no | yes | Subclade 2F |
| 2024-111 | Sep-2024 | Saxony-Anhalt | Hungary | yes | Subclade 2E |
| 2024-122 | Sep-2024 | Brandenburg | no | yes | Subclade 2E |
| 2024-124 | Sep-2024 | Schleswig-Holstein | unknown | yes | Subclade 2C |
| 2024-126 | Sep-2024 | Brandenburg | unknown | yes | Subclade 2F |
| 2024-130 | Sep-2024 | Saxony | unknown | yes | Subclade 2C |
| 2024-133 | Sep-2024 | Berlin | no | yes | Subclade 2C |

| | | | | | |
|---|---|---|---|---|---|
| 2024-135 | Sep-2024 | Saxony | unknown | no | n/a |
| 2024-141 | Sep-2024 | Lower Saxony | no | no | n/a |

**Supplementary Table S2. Reference dataset of West Nile virus complete or almost complete (≥70%) genome sequences with geographic and temporal metadata.** The table lists GenBank accession numbers, country and year of detection, and associated geographic coordinates (latitude and longitude) for each WNV strain.

| GenBank acc. No. | Country | Year of detection | Latitude | Longitude |
|---|---|---|---|---|
| DQ116961 | Hungary | 2004 | 46.3506900 | 20.1398580 |
| HQ537483 | Greece | 2010 | 40.8377778 | 22.8986111 |
| JN858070 | Italy | 2011 | 41.8933203 | 12.4829321 |
| KC407673 | Serbia | 2012 | 45.2551338 | 19.8451756 |
| KC496015 | Hungary | 2010 | 47.4979937 | 19.0403594 |
| KC496016 | Serbia | 2010 | 45.2551338 | 19.8451756 |
| KF179639 | Greece | 2012 | 40.9080556 | 24.6580556 |
| KF179640 | Austria | 2008 | 48.2083537 | 16.3725042 |
| KF588365 | Italy | 2013 | 44.9772062 | 12.2741904 |
| KF647248 | Italy | 2013 | 44.9772062 | 12.2741904 |
| KF647249 | Italy | 2013 | 44.9772062 | 12.2741904 |
| KF647250 | Italy | 2013 | 44.9772062 | 12.2741904 |
| KF647251 | Italy | 2013 | 45.4077172 | 11.8734455 |
| KF647252 | Italy | 2013 | 44.9772062 | 12.2741904 |
| KF823805 | Italy | 2013 | 44.9772062 | 12.2741904 |
| KF823806 | Italy | 2013 | 45.1692628 | 10.6708365 |
| KJ577738 | Greece | 2013 | 40.6569444 | 22.8894444 |
| KJ577739 | Greece | 2013 | 40.6769444 | 22.9100000 |
| KJ883341 | Greece | 2013 | 41.1300360 | 24.8864900 |
| KJ883342 | Greece | 2013 | 40.9822222 | 24.7047222 |
| KJ883343 | Greece | 2013 | 40.9813889 | 24.7994444 |
| KJ883344 | Greece | 2013 | 41.0627778 | 24.8969444 |
| KJ883345 | Greece | 2013 | 41.1300360 | 24.8864900 |
| KJ883346 | Greece | 2013 | 40.6216430 | 22.7528990 |
| KJ883347 | Greece | 2013 | 40.7047222 | 21.5219444 |
| KJ883348 | Greece | 2013 | 40.9822222 | 24.7047222 |
| KJ883349 | Greece | 2013 | 40.9822222 | 24.7047222 |
| KJ883350 | Greece | 2013 | 40.6105556 | 22.9755556 |
| KM203860 | Czech Republic | 2013 | 48.7825000 | 16.6702780 |
| KM203861 | Czech Republic | 2013 | 48.7825000 | 16.6702780 |
| KM203862 | Czech Republic | 2013 | 48.7886110 | 16.8172220 |
| KM203863 | Czech Republic | 2013 | 48.7886110 | 16.8172220 |
| KM659876 | Austria | 2014 | 48.2083537 | 16.3725042 |
| KP109691 | Austria | 2014 | 48.2083537 | 16.3725042 |
| KP109692 | Austria | 2014 | 48.2083537 | 16.3725042 |
| KP780837 | Austria | 2008 | 46.6785742 | 14.9069497 |

| | | | | |
|---|---|---|---|---|
| KP780838 | Austria | 2009 | 48.2083537 | 16.3725042 |
| KP780839 | Austria | 2011 | 48.2083537 | 16.3725042 |
| KP789953 | Italy | 2014 | 45.0368547 | 9.1378251 |
| KP789954 | Italy | 2014 | 45.2208641 | 10.0370380 |
| KP789955 | Italy | 2014 | 45.4384958 | 10.9924122 |
| KP789956 | Italy | 2014 | 45.4384958 | 10.9924122 |
| KP789957 | Italy | 2014 | 45.2208641 | 10.0370380 |
| KP789958 | Italy | 2014 | 45.0368547 | 9.1378251 |
| KP789959 | Italy | 2014 | 45.0368547 | 9.1378251 |
| KP789960 | Italy | 2013 | 45.1692628 | 10.6708365 |
| KT207792 | Italy | 2014 | 45.0696016 | 11.7715269 |
| KT359349 | Hungary | 2014 | 47.4979937 | 19.0403594 |
| KT757318 | Serbia | 2013 | 45.6012648 | 19.9715022 |
| KT757319 | Serbia | 2013 | 45.6012648 | 19.9715022 |
| KT757320 | Serbia | 2013 | 45.6012648 | 19.9715022 |
| KT757321 | Serbia | 2013 | 45.6012648 | 19.9715022 |
| KT757322 | Serbia | 2013 | 45.6012648 | 19.9715022 |
| KT757323 | Serbia | 2013 | 45.6012648 | 19.9715022 |
| KU206781 | Bulgaria | 2015 | 42.6983340 | 23.3199410 |
| KU573080 | Italy | 2013 | 44.8494142 | 11.6172101 |
| KU573081 | Italy | 2013 | 44.8494142 | 11.6172101 |
| KU573082 | Italy | 2013 | 44.8494142 | 11.6172101 |
| KU573083 | Italy | 2013 | 44.6501718 | 10.8867129 |
| KX375812 | Serbia | 2013 | 45.2551338 | 19.8451756 |
| KY594040 | Greece | 2010 | 40.6516667 | 23.3041667 |
| LR743421 | Germany | 2019 | 51.0508900 | 13.7383200 |
| LR743422 | Germany | 2019 | 52.9281500 | 12.8031100 |
| LR743423 | Germany | 2019 | 52.5029358 | 13.5205456 |
| LR743424 | Germany | 2019 | 52.5029358 | 13.5205456 |
| LR743425 | Germany | 2019 | 51.9064957 | 12.5417127 |
| LR743426 | Germany | 2019 | 52.5170365 | 13.3888599 |
| LR743427 | Germany | 2019 | 52.5170365 | 13.3888599 |
| LR743428 | Germany | 2019 | 52.5029358 | 13.5205456 |
| LR743429 | Germany | 2018 | 51.4062020 | 11.8889417 |
| LR743430 | Germany | 2019 | 51.0999765 | 13.6767995 |
| LR743431 | Germany | 2019 | 51.2701390 | 14.0948628 |
| LR743432 | Germany | 2019 | 52.5170365 | 13.3888599 |
| LR743433 | Germany | 2018 | 51.6796187 | 12.0611585 |
| LR743434 | Germany | 2018 | 48.1666670 | 11.8166670 |
| LR743435 | Germany | 2019 | 52.5873198 | 12.9920587 |
| LR743436 | Germany | 2018 | 51.5911653 | 12.5856428 |
| LR743437 | Germany | 2018 | 48.1667467 | 11.8037130 |

| | | | | |
|---|---|---|---|---|
| LR743442 | Germany | 2019 | 51.4825041 | 11.9705452 |
| LR743443 | Germany | 2018 | 52.5029358 | 13.5205456 |
| LR743444 | Germany | 2019 | 51.9064957 | 12.5417127 |
| LR743445 | Germany | 2019 | 50.8322608 | 12.9252977 |
| LR743446 | Germany | 2019 | 51.4825041 | 11.9705452 |
| LR743447 | Germany | 2019 | 52.5029358 | 13.5205456 |
| LR743448 | Germany | 2019 | 51.7567447 | 14.3357307 |
| LR743449 | Germany | 2019 | 51.6181969 | 12.2635906 |
| LR743450 | Germany | 2019 | 51.3406321 | 12.3747329 |
| LR743451 | Germany | 2019 | 52.1315889 | 11.6399609 |
| LR743452 | Germany | 2019 | 50.8322608 | 12.9252977 |
| LR743453 | Germany | 2019 | 51.4617493 | 12.4526970 |
| LR743454 | Germany | 2019 | 51.5911653 | 12.5856428 |
| LR743455 | Germany | 2019 | 52.5029358 | 13.5205456 |
| LR743456 | Germany | 2019 | 51.4390092 | 12.3714685 |
| LR743457 | Germany | 2019 | 51.3564413 | 11.9961480 |
| LR743458 | Germany | 2019 | 52.1315889 | 11.6399609 |
| LR989885 | Germany | 2019 | 52.5023038 | 13.5291672 |
| LR989888 | Germany | 2019 | 52.5023038 | 13.5291672 |
| MF984337 | Austria | 2015 | 48.2083537 | 16.3725042 |
| MF984338 | Austria | 2015 | 48.2083537 | 16.3725042 |
| MF984339 | Austria | 2015 | 48.2083537 | 16.3725042 |
| MF984340 | Austria | 2015 | 48.2083537 | 16.3725042 |
| MF984341 | Austria | 2015 | 48.1420000 | 16.1601000 |
| MF984342 | Austria | 2015 | 48.2083537 | 16.3725042 |
| MF984343 | Austria | 2015 | 48.2083537 | 16.3725042 |
| MF984344 | Austria | 2015 | 48.2083537 | 16.3725042 |
| MF984345 | Austria | 2015 | 48.2214000 | 16.2037000 |
| MF984346 | Austria | 2016 | 48.2083537 | 16.3725042 |
| MF984347 | Austria | 2016 | 48.2083537 | 16.3725042 |
| MF984348 | Austria | 2016 | 48.0040000 | 16.3417000 |
| MF984349 | Austria | 2016 | 48.2083537 | 16.3725042 |
| MF984350 | Austria | 2016 | 48.2083537 | 16.3725042 |
| MF984351 | Austria | 2016 | 48.2083537 | 16.3725042 |
| MF984352 | Austria | 2016 | 48.2083537 | 16.3725042 |
| MH021189 | Hungary | 2017 | 47.4811921 | 19.0602641 |
| MH244510 | Slovakia | 2014 | 48.1331000 | 17.2133000 |
| MH244511 | Slovakia | 2013 | 48.2131904 | 17.3603744 |
| MH244512 | Slovakia | 2013 | 48.2131904 | 17.3603744 |
| MH244513 | Slovakia | 2013 | 48.7015059 | 21.6555995 |
| MH549209 | Greece | 2017 | 37.6333306 | 22.7333306 |
| MH924836 | Germany | 2018 | 51.2924000 | 11.5822000 |

| | | | | |
|---|---|---|---|---|
| MH986055 | Germany | 2018 | 52.3219000 | 13.2137000 |
| MH986056 | Germany | 2018 | 54.0528000 | 12.0636000 |
| MK473443 | Greece | 2017 | 37.6333306 | 22.7333306 |
| MK947396 | Slovenia | 2018 | 46.0499803 | 14.5068602 |
| MK947397 | Slovenia | 2018 | 46.0499803 | 14.5068602 |
| MN480792 | Greece | 2018 | 40.6711111 | 22.9275000 |
| MN480793 | Greece | 2018 | 40.8182490 | 22.1837890 |
| MN480794 | Greece | 2018 | 40.7144444 | 23.0416667 |
| MN480795 | Greece | 2018 | 40.8227778 | 22.2491667 |
| MN481589 | Greece | 2018 | 40.5233333 | 23.0477778 |
| MN481590 | Greece | 2018 | 40.6216430 | 22.7528990 |
| MN481591 | Greece | 2018 | 41.0833330 | 25.4166670 |
| MN481592 | Greece | 2012 | 40.6216430 | 22.7528990 |
| MN481593 | Greece | 2012 | 40.6403167 | 22.9352716 |
| MN481594 | Greece | 2012 | 40.7947222 | 22.4116667 |
| MN481595 | Greece | 2010 | 40.6216430 | 22.7528990 |
| MN481596 | Greece | 2011 | 40.7947222 | 22.4116667 |
| MN481597 | Greece | 2012 | 40.7947222 | 22.4116667 |
| MN652878 | Greece | 2018 | 40.6216430 | 22.7528990 |
| MN652879 | Greece | 2018 | 40.8182490 | 22.1837890 |
| MN652880 | Greece | 2018 | 40.6140650 | 22.9783660 |
| MN794935 | Germany | 2019 | 51.1831000 | 12.2130000 |
| MN794937 | Germany | 2019 | 52.0121000 | 11.4429000 |
| MN794938 | Germany | 2019 | 51.4053000 | 12.1827000 |
| MN794939 | Germany | 2019 | 53.3012000 | 10.0314000 |
| MN939557 | Italy | 2018 | 44.9772062 | 12.2741904 |
| MN939558 | Italy | 2016 | 44.9772062 | 12.2741904 |
| MN939559 | Italy | 2016 | 44.9772062 | 12.2741904 |
| MN939560 | Italy | 2016 | 45.6348591 | 11.4063543 |
| MN939561 | Italy | 2018 | 45.4371908 | 12.3345898 |
| MN939562 | Italy | 2018 | 45.8066914 | 12.2063158 |
| MN939563 | Italy | 2018 | 45.4384958 | 10.9924122 |
| MN939564 | Italy | 2016 | 44.9772062 | 12.2741904 |
| MT341470 | Greece | 2019 | 39.7582800 | 22.3729700 |
| MT341471 | Greece | 2019 | 39.3978340 | 22.0700870 |
| MT341472 | Bulgaria | 2018 | 43.4090220 | 24.6180123 |
| MT863560 | France | 2018 | 48.8588897 | 2.3200410 |
| MT863561 | France | 2018 | 48.8588897 | 2.3200410 |
| MW036633 | Netherlands | 2020 | 52.1342798 | 5.0064058 |
| MW036634 | Netherlands | 2020 | 52.1357615 | 5.0034393 |
| MW142223 | Germany | 2020 | 51.3406321 | 12.3747329 |
| MW142224 | Germany | 2020 | 51.3406321 | 12.3747329 |

| | | | | |
|---|---|---|---|---|
| MW142225 | Germany | 2020 | 51.3406321 | 12.3747329 |
| MW142226 | Germany | 2020 | 51.3406321 | 12.3747329 |
| MW142227 | Germany | 2020 | 51.3406321 | 12.3747329 |
| MW561633 | Slovakia | 2018 | 48.7748988 | 21.2067674 |
| MW751834 | Serbia | 2018 | 45.4163890 | 19.8916670 |
| MW751835 | Serbia | 2018 | 46.0969440 | 19.6575000 |
| MW751836 | Serbia | 2015 | 45.7094440 | 19.6605560 |
| MW751837 | Serbia | 2018 | 45.4708330 | 20.0641670 |
| MW751838 | Serbia | 2018 | 44.8125000 | 20.4611110 |
| MW751839 | Serbia | 2020 | 45.2394440 | 19.8227780 |
| MW751840 | Serbia | 2020 | 45.2394440 | 19.8227780 |
| MW751841 | Serbia | 2018 | 45.5961110 | 20.1344440 |
| MW751842 | Serbia | 2018 | 46.0969440 | 19.6575000 |
| MW751843 | Serbia | 2018 | 45.4986110 | 19.1972220 |
| MW751844 | Serbia | 2018 | 45.7730560 | 19.1152780 |
| MW751845 | Serbia | 2018 | 45.0522220 | 20.4302780 |
| MW751846 | Serbia | 2018 | 46.0969440 | 19.6575000 |
| MW862073 | Italy | 2012 | 40.0599068 | 8.7481167 |
| MW862074 | Italy | 2013 | 40.6783330 | 8.6180560 |
| MW862075 | Italy | 2015 | 44.5622220 | 11.0347220 |
| MW862076 | Italy | 2015 | 44.5622220 | 11.0347220 |
| MW862077 | Italy | 2015 | 44.5944500 | 11.0497900 |
| MW862078 | Italy | 2015 | 45.7872220 | 8.6197220 |
| MW862079 | Italy | 2015 | 45.0961110 | 8.5550000 |
| MW862080 | Italy | 2015 | 44.4006400 | 11.2517700 |
| MW862081 | Italy | 2015 | 44.8372220 | 11.0294440 |
| MW862082 | Italy | 2015 | 45.5136110 | 8.8516670 |
| MW862083 | Italy | 2015 | 40.7259250 | 8.5556830 |
| MW862084 | Italy | 2015 | 44.7247500 | 11.0958800 |
| MW862085 | Italy | 2015 | 44.7247500 | 11.0958800 |
| MW862086 | Italy | 2016 | 40.7039391 | 9.7052936 |
| MW862087 | Italy | 2017 | 44.3985400 | 11.5854600 |
| MW862088 | Italy | 2017 | 44.6181100 | 11.4725700 |
| MW862089 | Italy | 2017 | 45.3280560 | 9.4058330 |
| MW862090 | Italy | 2018 | 44.7658100 | 11.4869500 |
| MW862091 | Italy | 2018 | 44.8114700 | 10.8041400 |
| MW862092 | Italy | 2018 | 44.8434600 | 11.6086800 |
| MW862093 | Italy | 2018 | 44.3889300 | 11.3426200 |
| MW862094 | Italy | 2018 | 40.1716670 | 8.8561110 |
| MW862095 | Italy | 2018 | 39.8527780 | 9.0513890 |
| MW862096 | Italy | 2018 | 45.1920500 | 9.1591700 |
| MW862097 | Italy | 2018 | 39.7395200 | 9.1111400 |

| | | | | |
|---|---|---|---|---|
| MW862098 | Italy | 2018 | 40.5654100 | 8.3209200 |
| MW862099 | Italy | 2018 | 39.9302500 | 9.6702500 |
| MW862100 | Italy | 2018 | 40.5863000 | 9.0034000 |
| MW862101 | Italy | 2018 | 44.3889300 | 11.3426200 |
| MW862102 | Italy | 2019 | 45.5794400 | 12.3733300 |
| MW862103 | Italy | 2019 | 44.3830560 | 7.4361110 |
| MW862104 | Italy | 2019 | 44.3194440 | 7.8644440 |
| MW862105 | Italy | 2019 | 39.9302500 | 9.6702500 |
| MW862106 | Italy | 2019 | 44.3907100 | 7.5482800 |
| MW862107 | Italy | 2019 | 44.7283330 | 8.6902780 |
| MW862108 | Italy | 2020 | 45.9213600 | 8.5518300 |
| MZ190464 | Kosovo | 2018 | 42.4647220 | 21.4669440 |
| MZ190465 | Kosovo | 2018 | 42.6236110 | 20.8938890 |
| MZ190466 | Kosovo | 2018 | 42.6236110 | 20.8938890 |
| MZ190467 | Kosovo | 2018 | 42.6633330 | 21.1622220 |
| MZ268007 | Netherlands | 2020 | 52.1556593 | 4.9247740 |
| MZ605382 | Hungary | 2004 | 47.1801000 | 19.5039900 |
| MZ964751 | Germany | 2021 | 52.5200000 | 13.4000000 |
| MZ964752 | Germany | 2021 | 52.4750000 | 13.4275000 |
| MZ964753 | Germany | 2021 | 52.4750000 | 13.4275000 |
| OK129329 | Hungary | 2016 | 46.9325000 | 18.2416670 |
| OK129330 | Hungary | 2016 | 47.2152780 | 20.7572220 |
| OK129331 | Hungary | 2015 | 46.9227780 | 18.3469440 |
| OK129332 | Hungary | 2015 | 46.9616670 | 18.9355560 |
| OK129333 | Hungary | 2017 | 47.4450000 | 19.6908330 |
| OK129334 | Hungary | 2017 | 47.1801000 | 19.5039900 |
| OK129335 | Hungary | 2017 | 47.0172220 | 20.2808330 |
| OK129336 | Hungary | 2017 | 47.7502780 | 19.0788890 |
| OK239658 | Hungary | 2018 | 47.4433330 | 21.3963890 |
| OK239659 | Hungary | 2018 | 46.8961110 | 19.6894440 |
| OK239660 | Hungary | 2018 | 47.4977780 | 19.0400000 |
| OK239661 | Hungary | 2018 | 46.5683330 | 20.6544440 |
| OK239662 | Hungary | 2018 | 46.4352780 | 19.4833330 |
| OK239663 | Hungary | 2019 | 47.9494440 | 21.7241670 |
| OK239664 | Hungary | 2018 | 46.9616670 | 18.9355560 |
| OK239665 | Hungary | 2018 | 47.5286110 | 21.6252780 |
| OK239666 | Hungary | 2018 | 46.6733330 | 21.0875000 |
| OK239667 | Hungary | 2018 | 46.4902780 | 19.7366670 |
| OK239668 | Hungary | 2018 | 47.5000000 | 19.9061110 |
| OK239670 | Hungary | 2018 | 47.4433330 | 21.3963890 |
| OK239671 | Hungary | 2018 | 46.5911110 | 19.0563890 |
| OK239672 | Hungary | 2018 | 48.0355560 | 21.0647220 |

| | | | | |
|---|---|---|---|---|
| OL840871 | Greece | 2018 | 40.8016800 | 22.0439800 |
| OL840872 | Greece | 2018 | 40.5466667 | 23.0194444 |
| OL840873 | Greece | 2018 | 39.7166667 | 22.3958333 |
| OL840874 | Greece | 2019 | 40.2805590 | 22.5058400 |
| OL840875 | Greece | 2019 | 41.1166667 | 24.9125000 |
| OL840876 | Greece | 2019 | 41.1294444 | 25.0413889 |
| OL840877 | Greece | 2019 | 41.7063889 | 26.3013889 |
| OL840878 | Greece | 2019 | 40.8252778 | 22.3872222 |
| OL840879 | Greece | 2019 | 41.1224390 | 24.3296733 |
| OL840880 | Greece | 2020 | 39.2688889 | 21.8111111 |
| OL840881 | Greece | 2020 | 41.2077778 | 23.0963889 |
| OL840882 | Greece | 2020 | 40.7819444 | 22.2750000 |
| OL840883 | Greece | 2019 | 41.0099020 | 23.3709390 |
| OL840884 | Greece | 2019 | 40.9211650 | 22.7403330 |
| OL840885 | Greece | 2019 | 40.6601020 | 22.6327750 |
| OL840886 | Greece | 2020 | 41.2057960 | 23.0749210 |
| OL840887 | Greece | 2020 | 41.1423050 | 23.2131110 |
| OL840888 | Greece | 2020 | 39.8390171 | 22.5064489 |
| OL840889 | Greece | 2020 | 41.1666040 | 22.7954260 |
| OL840890 | Greece | 2020 | 41.0963889 | 23.5022222 |
| OL840891 | Greece | 2020 | 41.1760680 | 23.3917730 |
| OL840892 | Greece | 2020 | 41.1955556 | 23.3711111 |
| OL840893 | Greece | 2020 | 41.0247222 | 23.3633333 |
| OL840894 | Greece | 2021 | 40.6191910 | 22.6672150 |
| OL840895 | Greece | 2021 | 40.7066667 | 22.2650000 |
| OL840896 | Greece | 2021 | 41.1760680 | 23.3917730 |
| OL840897 | Greece | 2021 | 40.5597840 | 22.5922160 |
| OL840898 | Greece | 2021 | 40.7089140 | 22.7300240 |
| OL840899 | Greece | 2021 | 40.7831450 | 22.5904050 |
| OM037670 | Spain | 2017 | 41.7469750 | 0.5944720 |
| OM037671 | Spain | 2020 | 41.6701857 | 0.5381169 |
| OM037672 | Spain | 2020 | 41.1134734 | 1.2379323 |
| OM037673 | Spain | 2020 | 41.1134734 | 1.2379323 |
| ON032488 | Italy | 2020 | 45.0000000 | 8.0000000 |
| ON032489 | Italy | 2020 | 45.0000000 | 8.0000000 |
| ON032490 | Italy | 2021 | 45.0000000 | 8.0000000 |
| ON032491 | Italy | 2021 | 45.6667000 | 9.5000000 |
| ON032492 | Italy | 2021 | 44.7499970 | 11.0000000 |
| ON032493 | Italy | 2021 | 44.7499970 | 11.0000000 |
| ON032494 | Italy | 2021 | 40.0000000 | 9.0000000 |
| ON032495 | Italy | 2021 | 44.7499970 | 11.0000000 |
| ON032497 | Italy | 2021 | 44.7499970 | 11.0000000 |

| | | | | |
|---|---|---|---|---|
| ON032498 | Italy | 2022 | 43.0000000 | 12.5000000 |
| ON755223 | Netherlands | 2020 | 52.1019996 | 5.1741660 |
| ON813230 | Italy | 2021 | 44.7499970 | 11.0000000 |
| ON813231 | Italy | 2021 | 44.7499970 | 11.0000000 |
| ON813232 | Italy | 2021 | 40.0000000 | 9.0000000 |
| ON813233 | Italy | 2022 | 45.0000000 | 8.0000000 |
| OP009516 | Italy | 2021 | 45.4167000 | 11.0333000 |
| OP009517 | Italy | 2021 | 45.6690800 | 12.2361400 |
| OP179287 | Hungary | 2021 | 46.8636884 | 20.5535415 |
| OP179288 | Hungary | 2021 | 47.5341670 | 19.0525000 |
| OP561452 | Italy | 2019 | 45.4064823 | 11.8212057 |
| OP561453 | Italy | 2019 | 45.4384958 | 10.9924122 |
| OP561454 | Italy | 2019 | 45.4384958 | 10.9924122 |
| OP561455 | Italy | 2019 | 45.4384958 | 10.9924122 |
| OP561456 | Italy | 2019 | 45.6576180 | 12.2669080 |
| OP561457 | Italy | 2019 | 44.9772062 | 12.2741904 |
| OP561458 | Italy | 2019 | 45.4064823 | 11.8212057 |
| OP561459 | Italy | 2019 | 45.4064823 | 11.8212057 |
| OP734238 | Italy | 2022 | 44.7499970 | 11.0000000 |
| OP734239 | Italy | 2022 | 44.7499970 | 11.0000000 |
| OP734240 | Italy | 2022 | 45.0000000 | 8.0000000 |
| OP734241 | Italy | 2022 | 45.0000000 | 8.0000000 |
| OP734242 | Italy | 2022 | 44.7499970 | 11.0000000 |
| OP734243 | Italy | 2022 | 44.7499970 | 11.0000000 |
| OP734244 | Italy | 2022 | 45.5000000 | 11.7500000 |
| OP762592 | Netherlands | 2020 | 52.1352540 | 5.0026990 |
| OP762593 | Netherlands | 2020 | 52.1355130 | 5.0039880 |
| OP762594 | Netherlands | 2020 | 52.1364700 | 5.0030690 |
| OP762595 | Netherlands | 2020 | 52.1361694 | 5.0034938 |
| OP762596 | Netherlands | 2020 | 52.0955124 | 5.0125341 |
| OP762597 | Netherlands | 2020 | 52.1057167 | 5.0121598 |
| OP804520 | Poland | 2022 | 52.2297700 | 21.0117800 |
| OP810565 | Germany | 2021 | 52.5243700 | 13.4105300 |
| OP810566 | Germany | 2021 | 52.5243700 | 13.4105300 |
| OP810567 | Germany | 2021 | 52.5243700 | 13.4105300 |
| OP850024 | Italy | 2022 | 44.7499970 | 11.0000000 |
| OP850025 | Italy | 2022 | 45.0000000 | 8.0000000 |
| OP850026 | Italy | 2022 | 46.1666660 | 13.0000000 |
| OP850027 | Italy | 2021 | 45.0000000 | 8.0000000 |
| OP850028 | Italy | 2021 | 44.7499970 | 11.0000000 |
| OP850029 | Italy | 2021 | 44.7499970 | 11.0000000 |
| OQ053515 | Greece | 2022 | 40.4166650 | 23.4999980 |

| | | | | |
|---|---|---|---|---|
| OQ053516 | Greece | 2022 | 39.6243308 | 22.4203650 |
| OQ053517 | Greece | 2022 | 40.6366641 | 22.9421629 |
| OQ053518 | Greece | 2022 | 39.6243308 | 22.4203650 |
| OQ053519 | Greece | 2022 | 40.6366641 | 22.9421629 |
| OQ053520 | Greece | 2022 | 40.2499990 | 22.4166650 |
| OQ053521 | Greece | 2022 | 40.6366641 | 22.9421629 |
| OQ053522 | Greece | 2022 | 40.6366641 | 22.9421629 |
| OQ053523 | Greece | 2022 | 40.6366641 | 22.9421629 |
| OQ053524 | Greece | 2022 | 38.8999964 | 22.5333312 |
| OQ053525 | Greece | 2022 | 40.9833000 | 22.8667000 |
| OQ053526 | Greece | 2022 | 41.0900000 | 23.5500000 |
| OQ053527 | Greece | 2022 | 40.4166650 | 23.4999980 |
| OQ053528 | Greece | 2022 | 40.2499990 | 22.4166650 |
| OQ053529 | Greece | 2022 | 40.2499990 | 22.4166650 |
| OQ053530 | Greece | 2022 | 38.8999964 | 22.5333312 |
| OQ053531 | Greece | 2022 | 40.7609800 | 22.5255800 |
| OQ053532 | Greece | 2022 | 40.6366641 | 22.9421629 |
| OQ053533 | Greece | 2022 | 38.8999964 | 22.5333312 |
| OQ053534 | Greece | 2022 | 39.6243308 | 22.4203650 |
| OQ053535 | Greece | 2022 | 40.6366641 | 22.9421629 |
| OQ053536 | Greece | 2022 | 41.0900000 | 23.5500000 |
| OQ053537 | Greece | 2022 | 38.8999964 | 22.5333312 |
| OQ053538 | Greece | 2022 | 38.8999964 | 22.5333312 |
| OQ053539 | Greece | 2022 | 40.2499990 | 22.4166650 |
| OQ204314 | Italy | 2022 | 43.4167000 | 11.0000000 |
| OQ204315 | Italy | 2022 | 37.6230400 | 13.9345700 |
| OQ326499 | Germany | 2021 | 51.0210809 | 11.0259564 |
| OQ725906 | Germany | 2022 | 52.5243700 | 13.4105300 |
| OQ725907 | Germany | 2022 | 52.5243700 | 13.4105300 |
| OQ725908 | Germany | 2022 | 52.5243700 | 13.4105300 |
| OQ725909 | Germany | 2022 | 52.5243700 | 13.4105300 |
| OR091151 | Switzerland | 2022 | 45.9919483 | 8.9285680 |
| OR091152 | Switzerland | 2022 | 45.9919483 | 8.9285680 |
| OR091153 | Switzerland | 2022 | 45.9919483 | 8.9285680 |
| OR091154 | Switzerland | 2022 | 45.9919483 | 8.9285680 |
| OR091155 | Switzerland | 2022 | 45.9919483 | 8.9285680 |
| OR091156 | Switzerland | 2022 | 45.9919483 | 8.9285680 |
| OR091157 | Switzerland | 2022 | 45.9919483 | 8.9285680 |
| OR091158 | Switzerland | 2022 | 45.9919483 | 8.9285680 |
| OX442271 | Germany | 2020 | 51.3401990 | 12.3601030 |
| OX442272 | Germany | 2020 | 50.8831600 | 12.8703100 |
| OX442273 | Germany | 2020 | 51.5917400 | 12.5849200 |

| | | | | |
|---|---|---|---|---|
| OX442274 | Germany | 2020 | 51.5000000 | 12.0000000 |
| OX442275 | Germany | 2020 | 51.5000000 | 12.0000000 |
| OX442276 | Germany | 2020 | 51.3401990 | 12.3601030 |
| OX442277 | Germany | 2020 | 52.1277300 | 11.6291600 |
| OX442278 | Germany | 2020 | 51.4290752 | 12.1144393 |
| OX442279 | Germany | 2020 | 51.3401990 | 12.3601030 |
| OX442280 | Germany | 2020 | 52.5243700 | 13.4105300 |
| OX442281 | Germany | 2020 | 51.3967800 | 12.2214100 |
| OX442282 | Germany | 2020 | 52.5141000 | 13.5198000 |
| OX442283 | Germany | 2020 | 52.4571648 | 13.5089980 |
| OX442284 | Germany | 2020 | 50.9787000 | 11.0328300 |
| OX442285 | Germany | 2020 | 52.5499978 | 13.5499978 |
| OX442286 | Germany | 2020 | 50.8802900 | 12.0818700 |
| OX442287 | Germany | 2020 | 51.7946400 | 11.7401000 |
| OX442288 | Germany | 2020 | 51.0879910 | 11.6285628 |
| OX442289 | Germany | 2020 | 52.5166646 | 13.3666652 |
| OX442290 | Germany | 2020 | 51.5000000 | 12.0000000 |
| OX442291 | Germany | 2020 | 52.1277300 | 11.6291600 |
| OX442292 | Germany | 2020 | 52.4499982 | 13.5666644 |
| OX442293 | Germany | 2020 | 52.5141000 | 13.5198000 |
| OX442294 | Germany | 2020 | 52.5817191 | 13.5747926 |
| OX442295 | Germany | 2020 | 52.3127264 | 13.0260670 |
| OX442296 | Germany | 2020 | 52.5243700 | 13.4105300 |
| OX442297 | Germany | 2019 | 52.5243700 | 13.4105300 |
| OX442298 | Germany | 2020 | 52.5440890 | 13.2374310 |
| OX442299 | Germany | 2020 | 51.7354400 | 14.6397100 |
| OX442300 | Germany | 2020 | 52.5131000 | 13.5554000 |
| OX442301 | Germany | 2020 | 50.8802900 | 12.0818700 |
| OX442302 | Germany | 2020 | 52.4388946 | 13.2743077 |
| OX442303 | Germany | 2020 | 51.3401990 | 12.3601030 |
| OX442304 | Germany | 2020 | 50.9787000 | 11.0328300 |
| OX442305 | Germany | 2020 | 52.3581212 | 13.6536306 |
| OX442306 | Germany | 2020 | 52.5036647 | 13.6089976 |
| OX442307 | Germany | 2020 | 52.5666644 | 13.3333320 |
| OX442308 | Germany | 2020 | 50.9787000 | 11.0328300 |
| OX442309 | Germany | 2020 | 52.2381200 | 12.9714000 |
| OX442310 | Germany | 2018 | 51.4676610 | 13.6213034 |
| OX442311 | Germany | 2020 | 52.4333316 | 13.2499990 |
| OX442312 | Germany | 2020 | 52.5366645 | 13.2038325 |
| OX442313 | Germany | 2020 | 52.3956703 | 11.2968290 |
| OX442347 | Germany | 2020 | 52.4455000 | 13.5745500 |
| OX442348 | Germany | 2020 | 52.5434536 | 13.5886203 |

| | | | | |
|---|---|---|---|---|
| OX451204 | Germany | 2020 | 51.7946400 | 11.7401000 |
| PP104328 | Italy | 2023 | 44.7499970 | 11.0000000 |
| PP104329 | Italy | 2023 | 44.7499970 | 11.0000000 |
| PP104330 | Italy | 2023 | 45.6667000 | 9.5000000 |
| PP104331 | Italy | 2023 | 44.7499970 | 11.0000000 |
| PP104332 | Italy | 2023 | 44.7499970 | 11.0000000 |
| PP104333 | Italy | 2023 | 45.6667000 | 9.5000000 |
| PP104334 | Italy | 2023 | 45.6667000 | 9.5000000 |
| PP104335 | Italy | 2023 | 44.7499970 | 11.0000000 |
| PP104336 | Italy | 2023 | 45.6667000 | 9.5000000 |
| PP104337 | Italy | 2023 | 45.6667000 | 9.5000000 |
| PP104338 | Italy | 2023 | 44.7499970 | 11.0000000 |
| PP104339 | Italy | 2023 | 44.7499970 | 11.0000000 |
| PP104340 | Italy | 2023 | 42.6612300 | 13.6990100 |
| PP104341 | Italy | 2023 | 45.6667000 | 9.5000000 |
| PP104342 | Italy | 2023 | 45.6667000 | 9.5000000 |
| PP104343 | Italy | 2023 | 44.7499970 | 11.0000000 |
| PP104344 | Italy | 2023 | 40.0000000 | 9.0000000 |
| PP104345 | Italy | 2023 | 45.0000000 | 8.0000000 |
| PP104346 | Italy | 2023 | 45.6667000 | 9.5000000 |
| PP104347 | Italy | 2023 | 44.7499970 | 11.0000000 |
| PP104348 | Italy | 2023 | 44.7499970 | 11.0000000 |
| PP104349 | Italy | 2023 | 44.7499970 | 11.0000000 |
| PP104350 | Italy | 2023 | 44.7499970 | 11.0000000 |
| PP104351 | Italy | 2023 | 46.1666660 | 13.0000000 |
| PP104352 | Italy | 2023 | 44.7499970 | 11.0000000 |
| PP104353 | Italy | 2023 | 45.6667000 | 9.5000000 |
| PP104354 | Italy | 2023 | 45.5000000 | 11.7500000 |
| PP104355 | Italy | 2023 | 44.7499970 | 11.0000000 |
| PP104356 | Italy | 2023 | 44.7499970 | 11.0000000 |
| PP104357 | Italy | 2023 | 44.7499970 | 11.0000000 |
| PP104358 | Italy | 2023 | 45.6667000 | 9.5000000 |
| PP104359 | Italy | 2023 | 44.7499970 | 11.0000000 |
| PP104360 | Italy | 2023 | 44.7499970 | 11.0000000 |
| PP104361 | Italy | 2023 | 44.7499970 | 11.0000000 |
| PP104362 | Italy | 2023 | 44.7499970 | 11.0000000 |
| PP104363 | Italy | 2023 | 44.7499970 | 11.0000000 |
| PP104364 | Italy | 2023 | 45.6667000 | 9.5000000 |
| PP104365 | Italy | 2023 | 45.6667000 | 9.5000000 |
| PP104366 | Italy | 2023 | 44.7499970 | 11.0000000 |
| PP104367 | Italy | 2023 | 45.0000000 | 8.0000000 |
| PP104368 | Italy | 2023 | 44.7499970 | 11.0000000 |

| | | | | |
|---|---|---|---|---|
| PP104369 | Italy | 2023 | 45.5000000 | 11.7500000 |
| PP104370 | Italy | 2023 | 45.0000000 | 8.0000000 |
| PP104371 | Italy | 2023 | 44.7499970 | 11.0000000 |
| PP104372 | Italy | 2023 | 45.0000000 | 8.0000000 |
| PP104373 | Italy | 2023 | 45.5000000 | 11.7500000 |
| PP104374 | Italy | 2023 | 44.7499970 | 11.0000000 |
| PP104375 | Italy | 2023 | 45.5000000 | 11.7500000 |
| PP104376 | Italy | 2023 | 44.7499970 | 11.0000000 |
| PP104377 | Italy | 2023 | 44.7499970 | 11.0000000 |
| PP212878 | Hungary | 2023 | 47.7693355 | 21.8614121 |
| PP212879 | Hungary | 2023 | 47.4979130 | 19.0402360 |
| PP212880 | Hungary | 2023 | 46.2753260 | 19.8890062 |
| PP212881 | Hungary | 2023 | 46.2530000 | 20.1482400 |
| PP212882 | Hungary | 2023 | 47.4979130 | 19.0402360 |
| PP212883 | Hungary | 2023 | 46.3335502 | 20.2082846 |
| PP388974 | Italy | 2023 | 42.8333330 | 12.8333330 |
| PQ053322 | Serbia | 2022 | 44.8188000 | 20.4245000 |
| PQ053323 | Serbia | 2018 | 43.8682000 | 21.3209000 |
| PQ053324 | Serbia | 2018 | 44.8189200 | 20.4599800 |
| PQ053325 | Serbia | 2018 | 44.8189200 | 20.4599800 |
| PQ053326 | Serbia | 2022 | 44.8159000 | 20.4863000 |
| PQ053327 | Serbia | 2022 | 44.8159000 | 20.4863000 |
| PQ053328 | Serbia | 2018 | 44.8189200 | 20.4599800 |
| PQ053329 | Serbia | 2022 | 44.8687000 | 20.4755000 |
| PQ053330 | Serbia | 2022 | 44.6891000 | 21.0122000 |
| PQ053331 | Serbia | 2023 | 44.4943570 | 21.0105533 |
| PQ435199 | Italy | 2024 | 42.8333330 | 12.8333330 |
| PQ435200 | Italy | 2024 | 42.8333330 | 12.8333330 |
| PQ435201 | Italy | 2024 | 42.8333330 | 12.8333330 |
| PQ435202 | Italy | 2024 | 42.8333330 | 12.8333330 |
| PQ435203 | Italy | 2024 | 42.8333330 | 12.8333330 |
| PQ435204 | Italy | 2024 | 42.8333330 | 12.8333330 |
| PQ435205 | Italy | 2024 | 42.8333330 | 12.8333330 |
| PQ462527 | Spain | 2024 | 41.6701857 | 0.5381169 |
| PV220993 | Germany | 2022 | 51.8956200 | 11.0562200 |
| PV220994 | Germany | 2023 | 51.8653729 | 12.6832710 |
| PV220995 | Germany | 2023 | 52.3751300 | 8.9653800 |
| PV220996 | Germany | 2024 | 51.4388605 | 14.2536419 |
| PV220997 | Germany | 2022 | 51.3396200 | 12.3712900 |
| PV220998 | Germany | 2023 | 51.7574200 | 11.4608400 |
| PV220999 | Germany | 2022 | 51.5000000 | 12.0000000 |
| PV221000 | Germany | 2022 | 49.5628172 | 10.0966496 |

| | | | | |
|---|---|---|---|---|
| PV221001 | Germany | 2022 | 51.35478 | 11.98923 |
| PV221002 | Germany | 2024 | 51.2285717 | 12.7961989 |
| PV221003 | Germany | 2024 | 48.3877501 | 11.2560622 |
| PV221004 | Germany | 2024 | 51.6666640 | 12.4333316 |
| PV221005 | Germany | 2024 | 51.6284100 | 12.2649200 |
| PV221006 | Germany | 2024 | 52.6078300 | 8.3700500 |
| PV221007 | Germany | 2024 | 52.6078300 | 8.3700500 |
| PV221008 | Germany | 2024 | 51.6537242 | 14.2236354 |
| PV221009 | Germany | 2024 | 52.6065900 | 12.3369600 |
| PV221010 | Germany | 2024 | 51.3396200 | 12.3712900 |
| PV221011 | Germany | 2024 | 50.9287800 | 11.5899000 |
| PV221012 | Germany | 2024 | 51.7116700 | 12.2753100 |
| PV221013 | Germany | 2024 | 52.4333330 | 10.9833330 |
| PV221014 | Germany | 2024 | 51.5925123 | 13.9245123 |
| PV221015 | Germany | 2024 | 53.8689300 | 10.6872900 |
| PV221016 | Germany | 2024 | 51.4742536 | 13.2692004 |
| PV221017 | Germany | 2024 | 51.2159083 | 12.3302888 |
| PV221018 | Germany | 2024 | 52.4393316 | 13.5013313 |
| PV221019 | Germany | 2020 | 51.1666670 | 13.5666670 |
| PV221020 | Germany | 2020 | 51.4615884 | 12.4531080 |
| PV221021 | Germany | 2020 | 51.0508900 | 13.7383200 |
| PV221022 | Germany | 2020 | 51.5000000 | 12.0000000 |

**Supplementary Table S3. Sequencing performance metrics for West Nile virus isolates (BNI-129, B956, UG37) during amplicon-seq optimization for WNV genome sequencing.** The table details Ct values estimated viral copy numbers (copies/µl) from serial dilutions; and sequencing metrics (read counts, merged/non-merged reads, discarded reads, clean reads, mapped and unmapped reads). Genome recovery percentages at 10x coverage are also provided.

| Sample ID | Dilution_factor | Ct_value | Copy/µl | Read_count | Merged_reads | Non_merged reads | Size dis-card_reads | Clean_reads | Mapped _reads | Unmapped_reads | Recovered Genome 10x |
|---|---|---|---|---|---|---|---|---|---|---|---|
| **WNV129-3** | $10^{-3}$ | 19.45 | 667225.7 | 398014 | 278982 | 119032 | 41502 | 237480 | 237402 | 78 | 100.00 |
| **WNV129-4** | $10^{-4}$ | 22.80 | 57291.35 | 383212 | 266980 | 116232 | 33628 | 233352 | 233212 | 140 | 100.00 |
| **WNV129-5** | $10^{-5}$ | 26.42 | 4036.214 | 341460 | 236586 | 104874 | 32570 | 204016 | 203914 | 102 | 100.00 |
| **WNV129-6** | $10^{-6}$ | 29.66 | 375.6640 | 331086 | 229552 | 101534 | 29600 | 199952 | 199826 | 126 | 98.61 |
| **WNV129-7** | $10^{-7}$ | 33.00 | 32.49365 | 361600 | 242288 | 119312 | 36490 | 205798 | 205786 | 12 | 95.71 |
| **WNV129-8** | $10^{-8}$ | 35.27 | 6.156583 | 200542 | 100830 | 99712 | 21706 | 79124 | 79108 | 16 | 33.8 |
| **WNVB956-2** | $10^{-2}$ | 21.54 | 144245.2 | 301962 | 213182 | 88780 | 26196 | 186986 | 186878 | 108 | 100.00 |
| **WNVB956-3** | $10^{-3}$ | 25.12 | 10464.46 | 287378 | 200688 | 86690 | 22022 | 178666 | 178588 | 78 | 100.00 |
| **WNVB956-4** | $10^{-4}$ | 28.66 | 781.7418 | 320080 | 224810 | 95270 | 26326 | 198484 | 198326 | 158 | 98.51 |
| **WNVB956-5** | $10^{-5}$ | 31.65 | 87.38857 | 291412 | 201860 | 89552 | 23416 | 178444 | 178188 | 256 | 96.65 |
| **WNVB956-6** | $10^{-6}$ | 34.77 | 8.881200 | 358804 | 239520 | 119284 | 62358 | 177162 | 177146 | 16 | 91.38 |
| **WNVB956-7** | $10^{-7}$ | 37.13 | 1.575321 | 205684 | 110802 | 94882 | 16942 | 93860 | 93848 | 12 | 32.96 |
| **WNVB956-8** | $10^{-8}$ | 40.52 | 0.131357 | 209176 | 64216 | 144960 | 33712 | 30504 | 30498 | 6 | 7.56 |
| **WNVUganda-1** | $10^{-1}$ | 19.73 | 543449.1 | 391444 | 280358 | 111086 | 42686 | 237672 | 237544 | 128 | 97.93 |
| **WNVUganda-2** | $10^{-2}$ | 23.17 | 43684.91 | 372114 | 262980 | 109134 | 39834 | 223146 | 223010 | 136 | 97.93 |
| **WNVUganda-3** | $10^{-3}$ | 26.80 | 3055.160 | 265138 | 189438 | 75700 | 28752 | 160686 | 160586 | 100 | 97.93 |
| **WNVUganda-4** | $10^{-4}$ | 29.86 | 324.4492 | 377430 | 263308 | 114122 | 40180 | 223128 | 223092 | 36 | 97.94 |
| **WNVUganda-5** | $10^{-5}$ | 32.81 | 37.34811 | 326524 | 223694 | 102830 | 37176 | 186518 | 186454 | 64 | 94.08 |
| **WNVUganda-6** | $10^{-6}$ | 36.79 | 2.021058 | 241750 | 129064 | 112686 | 49976 | 79088 | 79068 | 20 | 37.04 |
| **WNVUganda-7** | $10^{-7}$ | 39.13 | 0.363782 | 221104 | 126238 | 94866 | 12944 | 113294 | 113224 | 70 | 36.54 |

**Supplementary Table S4. Sequencing quality metrics for West Nile virus-positive blood donor samples analyzed by targeted amplicon sequencing and metagenomic sequencing.** The table details qRT-PCR results (Ct value, estimated copy number/µl); sequencing metrics (read counts, merged/non-merged reads, discarded reads, clean reads); and mapping results (mapped and unmapped reads). WNV genome recovery percentages at 1x and 10x coverage are also reported for each sample. Associated GenBank and SRA accession numbers for targeted amplicon sequences are provided. For samples additionally sequenced by metagenomics, read counts, genome recovery percentages, and corresponding SRA, BioProject, and BioSample accession numbers are listed.

| | | | Targeted_amplicon_sequencing | | | | | | | | | | | Metagenomic_sequencing | | | | | |
|---|---|---|---|---|---|---|---|---|---|---|---|---|---|---|---|---|---|---|---|
| Sample ID | Ct_value | Copy/µl | Read count | Merged reads | Non_merged reads | Size Discard reads | Clean reads | Mapped reads | Unmapped reads | Genome recovery (%) 1x | Genome recovery (%) 10x | Genbank accession | SRA accession | Read count | Genome recovery (%) 1x | Genome recovery (%) 10x | SRA accession | Bioproject ID | Biosample_ID |
| 20017945_2020 | 26.92 | 2797.965461 | 428608 | 376698 | 51910 | 29504 | 347194 | 346906 | 288 | 100 | 100 | PV221022 | SRR32328387 | 8088692 | 99.9537 | 99.8519 | SRR32330250 | PRJNA1223181 | SAMN46800585 |
| 11_2023 | 28.09 | 1187.062881 | 436402 | 349314 | 87088 | 43942 | 305372 | 300570 | 4802 | 100 | 97.77 | PV220998 | SRR32328418 | 15478394 | 72.8644 | 49.4771 | SRR32330281 | PRJNA1223181 | SAMN46800595 |
| 72_2024 | 29.26 | 503.6224722 | 908542 | 821390 | 87152 | 80502 | 740888 | 740698 | 190 | 100 | 100 | PV221007 | SRR32328405 | 11014758 | 98.8339 | 98.2323 | SRR32330268 | PRJNA1223181 | SAMN46800607 |
| 20015006_2020 | 29.29 | 492.6712132 | 419558 | 367464 | 52094 | 28820 | 338644 | 338534 | 110 | 100 | 97.62 | PV221020 | SRR32328424 | 23738990 | 99.6298 | 99.6298 | SRR32330287 | PRJNA1223181 | SAMN46800581 |
| 12_2022 | 29.94 | 305.9748491 | 612102 | 474146 | 137956 | 70062 | 404084 | 401998 | 2086 | 100 | 95.87 | PV220999 | SRR32328384 | 5910098 | 99.8056 | 98.8339 | SRR32330247 | PRJNA1223181 | SAMN46800588 |
| 04_2022 | 30.19 | 254.7532797 | 450954 | 390344 | 60610 | 27100 | 363244 | 363128 | 116 | 98.65 | 94.84 | PV220993 | SRR32328386 | 3578624 | 99.8889 | 99.5465 | SRR32330249 | PRJNA1223181 | SAMN46800586 |
| 0_2022 | 30.30 | 235.0232177 | 470324 | 399660 | 70664 | 33052 | 366608 | 365706 | 902 | 98.73 | 95.89 | PV221001 | SRR32328422 | 16602314 | 55.8908 | 49.8103 | SRR32330285 | PRJNA1223181 | SAMN46800591 |
| 04_2023 | 30.42 | 215.2380824 | 469442 | 402040 | 67402 | 33068 | 368972 | 367806 | 1166 | 97.77 | 95.87 | PV220994 | SRR32328421 | 11064566 | 89.218 | 84.137 | SRR32330284 | PRJNA1223181 | SAMN46800592 |
| 122_2024 | 31.15 | 126.0626853 | 545342 | 483132 | 62210 | 35872 | 447260 | 446010 | 1250 | 99.46 | 35096 | PV221014 | SRR32328395 | 5630374 | 99.7224 | 73.5308 | SRR32330258 | PRJNA1223181 | SAMN46800616 |
| 20015005_2020 | 31.40 | 104.9592233 | 371224 | 315680 | 55544 | 28432 | 287248 | 284928 | 2320 | 99.17 | 96.72 | PV221019 | SRR32328425 | 28487312 | 90.6155 | 83.9056 | SRR32330288 | PRJNA1223181 | SAMN46800580 |
| 126_2024 | 31.64 | 88.03133761 | 567386 | 497482 | 69904 | 44980 | 452502 | 439824 | 12678 | 99.33 | 93.68 | PV221016 | SRR32328393 | 4520072 | 1.6937 | 0 | SRR32330256 | PRJNA1223181 | SAMN46800618 |
| 20015957_2020 | 32.29 | 54.67211097 | 520998 | 401784 | 119214 | 31244 | 370540 | 370426 | 114 | 34182 | 90.57 | PV221021 | SRR32328391 | 28047092 | 81.8417 | 74.3174 | SRR32330254 | PRJNA1223181 | SAMN46800584 |
| 133_2024 | 32.68 | 41.08118554 | 591162 | 493128 | 98034 | 39402 | 453726 | 453520 | 206 | 94.98 | 86.61 | PV221018 | SRR32328390 | 4553876 | 27.1541 | 5.6085 | SRR32330253 | PRJNA1223181 | SAMN46800620 |
| 40_2024 | 32.90 | 34.96430792 | 607200 | 520838 | 86362 | 46846 | 473992 | 473926 | 66 | 100 | 97.81 | PV221004 | SRR32328412 | 12399722 | 89.9121 | 51.106 | SRR32330275 | PRJNA1223181 | SAMN46800600 |
| 54_2024 | 33.02 | 32.02088144 | 411722 | 364198 | 47524 | 34218 | 329980 | 329412 | 568 | 100 | 98.59 | PV221005 | SRR32328407 | 10820766 | 77.5567 | 22.6469 | SRR32330270 | PRJNA1223181 | SAMN46800605 |
| 130_2024 | 33.07 | 30.86882463 | 373580 | 325522 | 48058 | 21552 | 303970 | 301612 | 2358 | 97.35 | 85.87 | PV221017 | SRR32328392 | 5264460 | 31.2355 | 5.7103 | SRR32330255 | PRJNA1223181 | SAMN46800619 |
| 05_2024 | 33.29 | 26.27253998 | 467706 | 416950 | 50756 | 47146 | 369804 | 369646 | 158 | 99.19 | 35065 | PV220996 | SRR32328417 | 10456778 | 75.8723 | 12.2628 | SRR32330280 | PRJNA1223181 | SAMN46800596 |
| 111_2024 | 33.50 | 22.52509688 | 1965554 | 323776 | 1641778 | 150350 | 173426 | 173168 | 258 | 86.63 | 76.18 | PV221013 | SRR32328396 | 3423184 | 35.5484 | 0 | SRR32330259 | PRJNA1223181 | SAMN46800615 |
| 58_2024 | 33.53 | 22.03528916 | 601720 | 514320 | 87400 | 80172 | 434148 | 433908 | 240 | 35827 | 92.87 | PV221006 | SRR32328406 | 10286978 | 33.6974 | 2.397 | SRR32330269 | PRJNA1223181 | SAMN46800606 |
| 110_2024 | 33.90 | 16.80200675 | 2055456 | 171612 | 1883844 | 90380 | 81232 | 80966 | 266 | 75.68 | 63.46 | PV221012 | SRR32328397 | 3234628 | 7.2929 | 0 | SRR32330260 | PRJNA1223181 | SAMN46800614 |
| 97_2024 | 34.62 | 9.913139833 | 510504 | 445878 | 64626 | 36952 | 408926 | 408672 | 254 | 35947 | 95.21 | PV221010 | SRR32328400 | 7985194 | 77.242 | 17.131 | SRR32330263 | PRJNA1223181 | SAMN46800611 |
| 75_2024 | 34.64 | 9.768906585 | 561238 | 498366 | 62872 | 44742 | 453624 | 453280 | 344 | 100 | 36100 | PV221008 | SRR32328404 | 9768698 | 92.3276 | 65.6178 | SRR32330267 | PRJNA1223181 | SAMN46800608 |
| 124_2024 | 34.73 | 9.145389703 | 501206 | 433698 | 67508 | 17534 | 416164 | 415446 | 718 | 94.16 | 30164 | PV221015 | SRR32328394 | 4615674 | 15.3447 | 2.4248 | SRR32330257 | PRJNA1223181 | SAMN46800617 |
| 141_2024 | 34.78 | 8.816354149 | 2329404 | 747548 | 1581856 | 420170 | 327378 | 310002 | 17376 | 70 | 58.57 | na | SRR32328388 | 4631926 | 10.4859 | 0.472 | SRR32330251 | PRJNA1223181 | SAMN46800622 |
| 39_2024 | 34.84 | 8.437100074 | 596666 | 499628 | 97038 | 39504 | 460124 | 460076 | 48 | 28550 | 75.23 | PV221003 | SRR32328414 | 12738614 | 12.4942 | 0.1666 | SRR32330277 | PRJNA1223181 | SAMN46800599 |
| 18_2022 | 34.95 | 7.783665866 | 411570 | 297238 | 114332 | 25054 | 272184 | 271902 | 282 | 88.66 | 79.87 | PV221000 | SRR32328383 | 6228896 | 20.1296 | 12.8829 | SRR32330246 | PRJNA1223181 | SAMN46800589 |
| 81_2024 | 35.12 | 6.871939726 | 519684 | 440572 | 79112 | 55600 | 384972 | 384906 | 66 | 35217 | 89.84 | PV221009 | SRR32328403 | 8894960 | 24.9236 | 4.174 | SRR32330266 | PRJNA1223181 | SAMN46800609 |
| 20015175-33_2020 | 35.31 | 5.978733883 | 566582 | 119848 | 446734 | 112110 | 7738 | 7560 | 178 | 9.97 | 2.44 | na | SRR32328413 | 20890384 | 3.0634 | 0 | SRR32330276 | PRJNA1223181 | SAMN46800582 |
| 09_2023 | 35.39 | 5.638299231 | 425384 | 289058 | 136326 | 44290 | 244768 | 240394 | 4374 | 19480 | 46.53 | na | SRR32328419 | 12905976 | 1.5086 | 0 | SRR32330282 | PRJNA1223181 | SAMN46800594 |
| 92_2024 | 35.40 | 5.597131144 | 1205690 | 698020 | 507670 | 211892 | 486128 | 485918 | 210 | 11.92 | 45927 | na | SRR32328401 | 8295034 | 3.1282 | 0 | SRR32330264 | PRJNA1223181 | SAMN46800610 |
| 05_2023 | 35.44 | 5.435442822 | 303730 | 77666 | 226064 | 53794 | 23872 | 23120 | 752 | 49.46 | 39.86 | PV220995 | SRR32328420 | 11211908 | 4.9329 | 4.7478 | SRR32330283 | PRJNA1223181 | SAMN46800593 |
| 135_2024 | 35.53 | 5.088516549 | 865366 | 474808 | 390558 | 390214 | 84594 | 3242 | 81352 | 10.81 | 45933 | na | SRR32328389 | 5368130 | 0 | 0 | SRR32330252 | PRJNA1223181 | SAMN46800621 |
| 26_2024 | 35.55 | 5.01448014 | 632712 | 255296 | 377416 | 177582 | 77714 | 76704 | 1010 | 47.72 | 45.71 | na | SRR32328416 | 11284282 | 3.6835 | 0 | SRR32330279 | PRJNA1223181 | SAMN46800597 |
| 99_2024 | 35.58 | 4.905440385 | 2779154 | 210272 | 2568882 | 142602 | 67670 | 67382 | 288 | 57.24 | 48.46 | na | SRR32328399 | 4770488 | 12.522 | 0.7774 | SRR32330262 | PRJNA1223181 | SAMN46800612 |
| 100_2024 | 35.62 | 4.763733427 | 2394024 | 168866 | 2225158 | 93076 | 75790 | 75380 | 410 | 26755 | 24108 | PV221011 | SRR32328398 | 2780204 | 22.5174 | 0 | SRR32330261 | PRJNA1223181 | SAMN46800613 |
| 20015336-32_2020 | 35.95 | 3.74042028 | 437208 | 320770 | 116438 | 22390 | 298380 | 296854 | 1526 | 26.86 | 21.84 | na | SRR32328402 | 40852404 | 37.1957 | 30.8191 | SRR32330265 | PRJNA1223181 | SAMN46800583 |
| 37_2024 | 36.02 | 3.55338233 | 879910 | 526066 | 353844 | 208266 | 317800 | 315710 | 2090 | 72.59 | 25934 | PV221002 | SRR32328415 | 13773484 | 3.2207 | 0.7219 | SRR32330278 | PRJNA1223181 | SAMN46800598 |
| 19_2022 | 36.35 | 2.790068658 | 593338 | 410466 | 182872 | 42806 | 367660 | 367286 | 374 | 56.76 | 18872 | na | SRR32328423 | 4163236 | 27.6724 | 26.2286 | SRR32330286 | PRJNA1223181 | SAMN46800590 |
| 51_2024 | 36.40 | 2.689686737 | 683000 | 302696 | 380304 | 193802 | 108894 | 104538 | 4356 | 23.58 | 22.54 | na | SRR32328409 | 11082048 | 0 | 0 | SRR32330272 | PRJNA1223181 | SAMN46800603 |
| 49_2024 | 36.98 | 1.758363695 | 490972 | 207604 | 283368 | 117524 | 90080 | 89780 | 300 | 39.48 | 13271 | na | SRR32328411 | 11541178 | 5.2661 | 0 | SRR32330274 | PRJNA1223181 | SAMN46800601 |
| 50_2024 | 37.98 | 0.844977076 | 773080 | 333678 | 439402 | 245692 | 87986 | 86706 | 1280 | 13.55 | 5.34 | na | SRR32328410 | 11896144 | 0 | 0 | SRR32330273 | PRJNA1223181 | SAMN46800602 |
| 07_2022 | 39.00 | 0.400143595 | 554550 | 409558 | 144992 | 31958 | 377600 | 377512 | 88 | 64.27 | 58.52 | PV220997 | SRR32328385 | 4304072 | 16.261 | 7.2837 | SRR32330248 | PRJNA1223181 | SAMN46800587 |
| 52_2024 | 43.63 | 0.013447903 | 897692 | 566432 | 331260 | 157692 | 408740 | 408482 | 258 | 44.37 | 42.75 | na | SRR32328408 | 12200048 | 0 | 0 | SRR32330271 | PRJNA1223181 | SAMN46800604 |
